# The Effectiveness of Albendazole in Reducing Soil-Transmitted Helminth Infection Among 9 to 12 year-old Students from 5^th^ Grade of a Public Elementary School in the Philippines

**DOI:** 10.1101/2025.05.07.25326997

**Authors:** Dihiansan Joan Christy, Elumba Erikka, Acebes Ma. Jobelle, Aguhayon Pinky Rose, Aluben Angelica Louise, Cabalhin Sharyz Dane, Chandrasekaran Rajakumar, Galindo Lemuel Hope, Talili Giovanni Sergius

**Affiliations:** Cebu Institute of Medicine

**Keywords:** Soil-Transmitted Helminths, DOH school-based national deworming program, CR, ERR, grade five students, Albendazole

## Abstract

**Context:** Soil-Transmitted Helminth (STH) infections are among the neglected tropical diseases with the highest prevalence and incidence in children aged 6 to 15 years. Appropriate health and nutrition interventions are necessary to prevent long-term adverse effects of parasitic infection. One example is the biannual school-based mass drug administration by the Department of Health in partnership with the Department of Education, as recommended by the World Health Organization.

**Objectives:** The general objective of the study is to determine the effectiveness of albendazole in reducing STH infection among school-aged children in 5th grade. Specifically, the study aims to identify the most common type of STH infection among the respondents, and to calculate the cure rate (CR) and egg reduction rate (ERR) overall and for each helminth species.

**Study Design:** Descriptive and comparative

**Study Setting:** Stool samples from the students were collected at a local public elementary school and processed in the Microbiology Laboratory of the Cebu Institute of Medicine.

**Participants:** All grade five students were invited to participate, with a final sample size of 188 respondents.

**Maneuver:** A total of four stool samples were collected from each participant: two before deworming (July 16-26, 2019) and two more 14 to 21 days after the first deworming cycle. All samples were analyzed using the Kato-Katz method. McNemars test was used as the statistical tool.

**Main Outcome Measures:** Percentage and intensity of STH infection, cure rate (CR), egg reduction rate (ERR), and effectiveness of intervention

**Results:** Of the 49 respondents, 10 (20.41%) were positive for STH at baseline. Single infection with Ascaris lumbricoides (12.24%) was most frequent, followed by co-infection with A. lumbricoides and Trichuris trichiura (4.08%), and single infection with T. trichiura (2.04%). Albendazole yielded a 100% CR and ERR for A. lumbricoides. CR and ERR for T. trichiura were 33.33% and 51.25%, respectively.

**Implications:** The drug is effective in reducing viable egg count in infected patients but is not sufficient to cure patients with T. trichiura infection.

**Conclusion:** A single 400 mg dose of Albendazole was effective against ascariasis. It was also effective against trichuriasis in terms of egg reduction, but not in terms of cure rate.

**Recommendations:** The DOH protocol should be re-evaluated regarding the use of a triple dose of albendazole instead of a single dose to address T. trichiura infection. Follow-up surveillance is recommended to monitor the effectiveness of the deworming program. Educating stakeholders to increase awareness and participation is also advised. Future researchers should investigate possible risk factors contributing to STH infection to inform further preventive action.

## Introduction

### Background of the Study

Soil-Transmitted Helminthiases, also known as intestinal worm infections, is a condition in which a parasite infects the gastro-intestinal tract of humans and other animals.^1^ According to the World Health Organization, the whole Philippine archipelago is endemic of STH.

In line with this, the Philippine Department of Health (DOH), in partnership with the Department of Education (DepEd), implemented the Integrated Helminth Control Program (IHCP) in 2006. The program aimed to address the prevalence of soil-transmitted helminthiases through deworming of at least 85% of the children age 1 to 12 years old and decrease the prevalence of STH infections among adolescent females, pregnant women and special population groups to less than 50% by the year 2010. School-age children (SAC) aged 6 to 15 years old are the primary target of WHO since this age group is observed to have the highest prevalence and intensity level of STH infections compared to the other age groups.^2^

The World Health Organization (WHO) recommends that the deworming process should take place twice a year to prevent the return to the original pre-treatment levels of infection.^3^ A school-based mass drug administration (MDA) for children in the Philippines is conducted twice a year for three consecutive years followed by a yearly MDA. The IHCP also encompasses the other factors that contribute to the pervasiveness of STH infections such as water, sanitation, environment, snail control, hygiene, and health education.^4^

However, despite the implementation of the IHCP in various municipalities in the Philippines, STH infections continue to persist in several provinces in the Philippines. An assessment of several municipalities in Masbate revealed that the infection rates and intensity of STH remained high and has failed to meet the IHCP and WHO reduction targets after a decade from the implementation of the IHCP.^5^

Moreover, in 2013, the Director of the National Institute of Health in the Philippines questioned the effectiveness of the program because it only covered 20% of affected children with an infection rate of 44%. This may be due to the fact that the protocol is only partially effective in achieving morbidity control; however, it does not prevent re-infection. This means that once treatment is stopped, prevalence returns to pre-treatment levels within 12–18 months.^6, 7, 8^ Therefore, it is recommended to use a combination of interventions that prevent re-infection alongside those that boost immunity (e.g., the use of micro/macronutrient supplements) in order to enhance the effectiveness of the treatment.

Intestinal helminths are endemic throughout the Philippines and efforts are underway to decrease their burden. However, there is limited evidence with regards to their prevalence, intensity and their impact on children’s nutritional status. As such, the purpose of this study is to determine the prevalence and intensity of STH infection in a public elementary school in the Philippines and examine the relationship between level of awareness, sanitation practices, and other identified risk factors associated with STH infections.

### Significance of the Study

Studies regarding the effectiveness of the deworming program in the Philippines are still of limited resource since the DOH does not conduct an annual assessment of the program itself. The present study examines the effectiveness of the drug used, Albendazole, by the current deworming program of the DOH – providing an evidence-based basis for the possible areas of improvement so as to further promote health and its accessibility.

The study benefits the students by encouraging them to develop proper hygienic practices improving their over-all well-being. Decreased transmission of STH infections leads to increased attendance and productivity of students in school.

The study benefits the parents by raising awareness regarding the health status, specifically the presence of STH infections, among their children so that they may teach and guide them to practice proper hygiene and help eliminate risk factors that may lead to STH infections. This will ultimately reduce unnecessary expenses that may be brought about by the hospitalization of children infected with STH.

The study benefits the community by giving them a sense of urgency on the need to provide ways on how to reduce the spread of STH infections by implementing proper sanitation in their environment, decreasing child mortality; thus, promoting a healthy and happy community.

The researchers can benefit through this study by gaining knowledge regarding the risk factors, adverse effects, prevalence, and effectivity of treatments associated with STH infections. This study will also provide additional information on the current status of the deworming program of the Department of Health.

## Objectives of the Study

### General Objective

To determine the effectiveness of Albendazole in reducing the prevalence and intensity of Soil-Transmitted Helminths among nine to twelve year-old students from grade five of a public Elementary School

### Specific Objectives

Specifically, this study aims to address the following:

1. To determine the demographic profile of the respondents when they are grouped according to :

- age
- sex
2. To determine what type of STH infection is the most common among the respondents.
3. To calculate the total cure rate (CR) and eggs reduction rate (ERR) along with CR and ERR for each species of the aforementioned soil transmitted helminths.

### Review of Related Literature

The present literature review is divided into four sections. The first section provides an overview of previous similar studies that provides an overview on soil-transmitted helminths, per se, and its prevalence of STH infections in both national and global scale. On the other hand, the second section discusses the clinical correlates of the infection caused by soil-transmitted helminths. The third section reviews the current preventive and treatment regimen to STH infection. Finally, the fourth section outlines the deworming program of the government, as measure against the increasing incidence of STH infections in the country.

### Overview on Soil-transmitted Helminths (STHs)

Human soil-transmitted helminths (STHs) include: (a) *Ascaris lumbricoides*, *Trichuris trichiura*, and the hookworms such as *Necator americanus* and *Ancylostoma duodenale*.^9, 10^ STH infections are transmitted via exposure to soil or ingestion of the products thereof (e.g. raw agricultural foods), which is contaminated with the eggs of these parasites. These eggs are present in human feces—thus, it is most likely to contaminate the soil and water in areas with sub-optimal systems of sanitation.^11^ In connection, a higher incidence of STHs incidence has also been observed among individuals from lower socioeconomic status. On the other hand, exposure to the contaminated soil could also involve playing barefoot on these areas, which makes children with unsafe hygiene practices vulnerable to these infections. ^1^

*Ascaris lumbricoides* is commonly known as the large intestinal roundworm of man. Ascariasis, the infection caused by *Ascaris lumbricoides*, begins following the ingestion of infective eggs containing larvae. In the small intestine, the larvae emerge from the eggs, and undergoes a liver-lung migration by entering the bloodstream via intestinal wall penetration. Once inside the lung, they migrate into the bronchioles, and are coughed up into the pharynx. They are then swallowed back into the small intestine. Maturation of larvae into adult worms occurs in the small intestine. In here, female worms produce approximately 200,000 eggs per day which are passed in the feces. Eggs embryonate and become infective in the outside environment especially in warm and moist soil. The people who are most susceptible to harbor Ascariasis are those who live in regions with warm climates and areas of poor sanitation particularly where human feces is used as a fertilizer, and where people directly defecate on the ground. The population most at risk are children who place their contaminated hands into their mouths.^12^

*Trichuris trichiura*, commonly known as whipworm, causes Trichuriasis. Infection is initiated by the ingestion of infective eggs containing larvae. The larvae emerge from the eggs in the small intestine. They grow and develop within the intestinal villi, and migrate into the cecum where they undergo complete maturation. Adult worms reside in the cecum and ascending colon. Female worms lay approximately 3,000-20,000 undeveloped eggs per day which are passed in the feces. Eggs develop and become infective in the soil. Infections with both *Trichuris trichiura* and *Ascaris lumbricoides* commonly occur since the mode of transmission is identical for both parasites.^13^

*Necator americanus* and *Ancylostoma duodenale* are commonly known as the new world hookworm and old world hookworm, respectively. Hookworm infection, either Ancylostomiasis or Necatoriasis begins when the third-stage filariform larvae, the infective stage of hookworm, penetrates the skin particularly unprotected feet. They migrate into the lymphatics and bloodstream, and undergo lung migration. They penetrate the capillaries, and enter the alveoli and bronchioles. They are coughed up into the pharynx and are swallowed into the small intestine, where they mature, live and multiply. Adult worms attach to the intestinal walls which results to blood loss by the host. Eggs laid by female worms are passed in the feces. The first stage rhabditiform larvae emerge from the eggs in the soil. They develop into the infective third-stage filariform larvae by molting twice.^14^

### Prevalence

A study conducted in ten randomly selected elementary schools in Tublay, Benguet, Philippines showed that out of the 428 students tested, only 2.34% of the respondents presented with STH infections. There was a significant correlation between the prevalence rate of soil-transmitted infections and factors such as: extent of knowledge on STH, practices on hand washing and on wearing of footwear of the respondents. No significant correlation; however, was found between the prevalence rate and the type of toilet used, and water source of the respondents. This shows that students who lacked awareness regarding STH infection and transmission, as well as proper health and sanitation practices, had a higher rate of getting infected.^15^ A meta-analysis of related literature by Addiss et al. supported these findings by associating access to sanitation with a lower chance of infection with any STH, *T. trichiura* and *A. lumbricoides* but not with hookworm infection, while wearing of shoes was the primary factor in decreasing the incidence of acquiring hookworm infection since its major mode of transmission is through skin penetration of mature larvae from hookworm eggs that hatched in the soil. Handwashing with soap prior to and after eating and defecation also significantly reduced infection with STH. ^16, 17^

Monitoring of the school-based helminth control program that was carried out by a group of researchers from Cavite Province, Philippines noted that although there was a significant decrease in the overall cumulative and high intensity prevalence of STH infections from baseline after mass treatment coverage by the government, the results remained high with regards to the standards set by the IHCP (close to 50% and at 24.6%, respectively). One remarkable finding was that undernourished students had the highest cumulative and high intensity prevalence of STH infections whereas students from the school district with the highest overall NAT scores also had the lowest rates of undernourished and STH infected children both at baseline and follow-up.^18^

In a cross-sectional survey carried out in Northern Samar, the Philippines, moderate to high prevalence rates of *A. lumbricoides (*36.5%), *T. trichiura (*61.8%), hookworm (28.4%), and harboring any of the aforementioned helminths (75.6%) were found among the residents in 18 rural barangays where majority of the villagers were poor rice farmers. The major risk factors for acquiring STH infections were identified as: low socio-economic status, low educational attainment, poor personal hygiene, frequency of water contact in rivers/lakes, occupation, and male gender.^19^

A study by Roque et. al (unpublished data, 2014) which included a total of 67 students aged 10 to 12 years old from Grades 4 to 6 of Banawa Elementary school showed that prevalence rates of infection were 4.35%, 36.36%, and 31.82%, respectively. Chi-square was computed and a significant correlation between the prevalence of Soil-Transmitted Helminthiasis and grade level was determined. According to gender, 68.75% of those positive were females, 31.25% were males. The percentage of those who were positive for Ascariasis was 93.75%, and those positive for Trichuriasis was 6.25%.

A similar study was done in 2015 by Abarquez, et al. (unpublished data, 2015) but more specifically aimed to assess the prevalence of Helminthiasis among Grade 4 students, within the age frame of 10-12 years old, of Lahug Elementary School. Only 436 out of 928 Grade 4 students were invited to participate as subjects. However, only 262 submitted stool samples; 109 of which were males and 153 were females. Samples were collected in batches during the month of February, and were examined through routine fecalysis and microscopic evaluation. An *Ascaris lumbricoides*, *Trichuris trichiura*, or *Hookworm spp.* seen, regardless of the stage [in the life cycle] it is in, was considered positive. Out of the 262 students, 8% were positive for STH infection. The prevalence of *Ascaris lumbricoides*, *Trichuris trichiura*, and *Hookworm spp.* respectively are as follows: 7.63%, 1.52%, and 0%. Most of those who were positive for the infection were males; however, there was no statistical significance [determined] between the two sexes. As mentioned, target sample size was not attained due to several factors identified by the researchers, such as the time frame, improper orientation and guidance, and the students’ lack of interest.

With the absence of a true gold standard in determination of soil transmitted helminths in fecal samples, the Kato-Katz (KK) thick smear technique has been recommended by the World Health Organization (WHO) for measuring its prevalence and monitoring the effectivity of control programs for moderate to high intensity cases (proportion of infected individuals is >20-50%) due to its cost-effective and quantitative nature; however, test sensitivity is reduced in low/light intensity infections (proportion of infected individuals is <20%) as the helminths could be overlooked because of uneven distribution of eggs within the rather small quantity of sample examined and because of rapid degradation and decline of hookworm eggs and larvae. As such, processing of at least two fresh stool samples collected in different days in duplicate readings is recommended to increase sensitivity and reduce diagnostic error. A study conducted about the sensitivity of Kato-katz for diagnosis of hookworm states that the sensitivity of the technique is dominated by high day-to-day variation and they recommend the collection of at least two samples over subsequent days. ^20, 21, 22, 23, 24, 25^ According to various studies, the FLOTAC technique is seen as a promising and more sensitive technique for helminth ova detection in low intensity STH infections, particularly for hookworm diagnosis. ^26, 27^ Drawbacks of the test include higher cost and sophistication of laboratory equipment to be used. ^28^

Other advancements in molecular based diagnostic tests with higher sensitivities than the KK technique are also continuously being developed. In Laguna Province, Philippines, a research carried out on STH infections among students showed that compared to the traditional Kato-Katz (KK) microscopic technique, the multiplex quantitative polymerase chain reaction (qPCR) method provided a higher degree of sensitivity for the detection of intestinal parasites: *Ascaris lumbricoides* (20.5% by KK vs. 60.8%hj by qPCR) and *Trichuris trichiura* (23.6% by KK vs. 38.8% by qPCR). Other parasitic species such as *Ancylostoma spp.*, *Necator americanus*, and *Strongyloides stercoralis*, which were not detected using the KK technique were identified using qPCR method.^1^ Recent studies also reinforce the prospect of using qPCR for quantitative STH assays owing to its superior diagnostic sensitivity and specificity for the aforementioned parasites especially for light intensity infections. ^29^ Despite the remarkable features of the qPCR assay, its high cost is the major hindrance to its utilization in most endemic regions where poverty and inadequacy of resources are rampant. ^30, 31^

### Clinical Correlates of the STH Infections

STH infections causes a number of symptoms and may lead to various complications when left untreated. The symptoms include general malaise, weakness and intestinal manifestations like diarrhea and abdominal pain.^32^ *A. lumbricoides* may lead to permanent growth impairment aside from acute infections like appendicitis, peritonitis, intestinal obstruction, and biliary disease.^33^ *T. trichiura* affects the mental growth and the children’s cognition as well as their general well-being.^34^ Growth retardation and inhibition of sexual development which are some of the characteristics of Crohn’s disease was attributed to Zinc deficiency, a deficiency appearing in intense *T. trichiura* infection.^35^ Zinc is considered a “type 2 nutrient” which means that there are no real stores for this. Low plasma value of zinc in STH infections or other conditions would then be caused by either low dietary intake of zinc of malnutrition, bearing in mind that poor nutritional status and STH infections are linked with each other. Malnutrition may increase susceptibility to STH infection, at the same time, STH infection could also lead to malnutrition.^36^ Hookworm infection may cause chronic intestinal blood loss leading to anemia specifically iron-deficiency anemia and albuminemia. ^37, 38^

### Preventive and Treatment Regimen to STH Infections

#### Effectiveness of Deworming Program

A cluster-randomized study, in 2013, was conducted in 72 administrative blocks in India with 2,576 6 to 72-month old children as its respondents. This particular study spanned 5 calendar year with 11 6-monthly mass treatment days. Their findings showed a decrease in nematode egg prevalence by 20% 1 to 5 months after the conduction of the albendazole deworming program.^39^

Another cluster-randomized controlled trial was conducted in Rural China which included 2,240 sample children, between the ages 9-11 years of age attending primary school for 2013-2014 school year, and spanned 112 townships in seven of the poorest rural counties in Qiandongnan Prefecture in Guizhou Province. They collected 2 fecal samples per child. This study used the Kato-Katz thick smear technique to test for *A. lumbricoides* (Ascaris), *T. trichiura* (Trichuris), and *A. duodenale* or *N. americanus* (hookworm). One smear was tested on the same day while the second smear was treated with formaldehyde and sent to the headquarters of the National Institute for Parasitic Diseases in Shanghai for quality control analysis and to perform egg counts for intensity of infection. The intervention used in this study was a distribution of a 400-mg albendazole dose accompanied by two educational pamphlets about STH infection, treatment, and prevention. Follow up surveys and fecal sample collection were performed in April 2014. The results of this particular study showed a 57.3% decrease in fecal egg counts in the intervention group while the control group experienced a 0.1% increase of mean fecal egg counts.^40^

#### Mebendazole and Albendazole

A meta-analysis was performed to assess the efficacy of mebendazole in six other countries Brazil, Cambodia, Cameroon, Ethiopia, United Republic of Tanzania, and Vietnam. This study also compared the FECR rate between mebendazole and albendazole. They found that mebendazole was most effective for *A. lumbricoides*, followed by hookworm then *T. trichiura*. When compared to albendazole, mebendazole was significantly more efficacious against hookworm and A. lumbricoides infections but equally efficacious for T. trichiura infections.^41^

Another study in Lao, randomly assigned 100 children to albendazole and another to mebendazole. After 21-23 days posttreatment, the cure rate and egg reduction rate showed that albendazole cured infection and reduced intensity of infection higher than mebendazole.^42^

#### Medication in DOH

The DOH uses chewable deworming pills of 400mg Albendazole or 500mg Mebendazole – antihelminthic drugs used for the treatment of a variety of parasitic worm infections against *Ascaris*, pinworm diseases, hookworm infections (*Ancylostoma duodenale*, *Necator americanus*), giardiasis, filariasis, trichuriasis (whipworm infections – *Trichuris trichiura*), neurocysticercosis, guinea worm disease and hydatid disease.

Albendazole has been found to be significantly most effective among other antihelminthic drugs like Pyrantel pamoate, Levamisole and Thiabendazole.^43, 44^ Albendazole has been found to be significantly more effective against hookworms than mebendazole, and a triple dose regimen of either drug is more effective than a single dose, but none is 100% effective all the time. Results of the test are in tabulated below.^45^

**Table 1.1.**
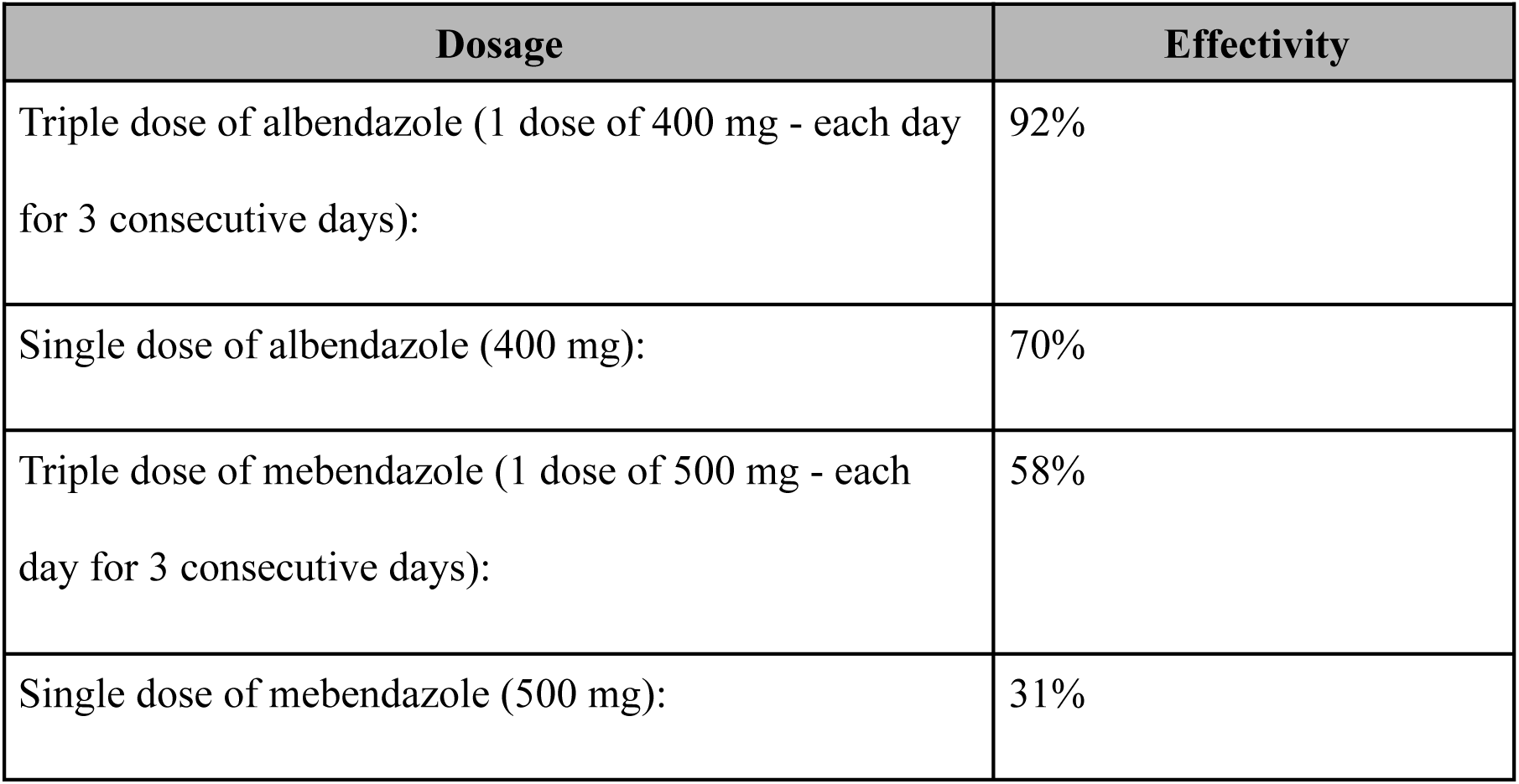
Results from a test of antihelminthic drugs against T. trichiura, A. lumbricoides and Taenia spp.

Single doses of anthelminthics are not very effective against Trichuris trichiura. For example, in one study, a single 500 mg dose of mebendazole only achieved a cure rate of 25.5%^46^

The US Centers for Disease Control and Prevention (CDC) state that whipworm is effectively treated using albendazole, mebendazole or ivermectin, but that each drug needs to be taken for 3 days^47^:

**Table 1.2.** Drug and each of their suggested dosage.

| Drug | Dosage for adults and children |
| --- | --- |
| Albendazole | 400 mg orally for 3 days |
| Mebendazole | 100 mg orally twice a day for 3 days. |
| Ivermectin | 200 mcg/kg/day orally for 3 days |

Although the CDC recommend the use of albendazole and mebendazole, these drugs have been shown to have low efficacy against *T. trichiura*, and their efficacy is continuing to weaken. It is also thought unlikely that these drugs will be effective against unhatched Trichuris eggs. A head-to-head comparison of three different drug combinations showed the highest efficacy was achieved by the combination of albendazole and oxantel pamoate^48^:

**Table 1.3.**
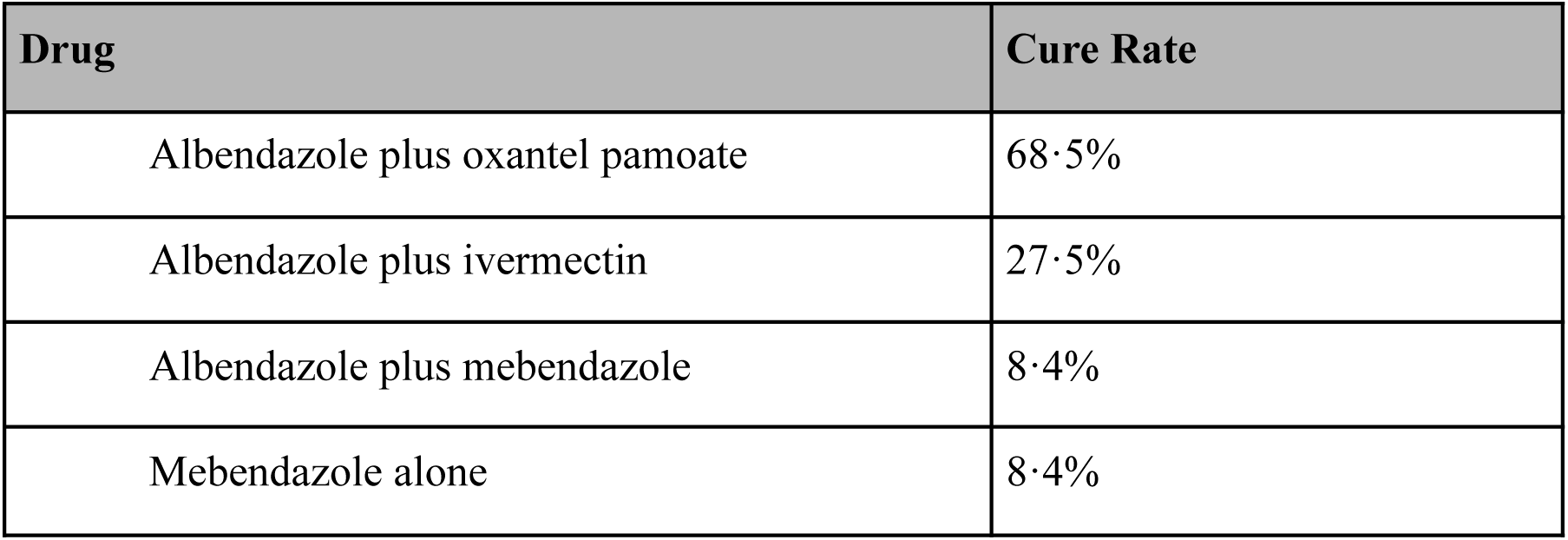
Drugs and each of their Cure Rates.

Oxantel pamoate may not be available, and mebendazole has been found to be marginally more effective against whipworm than albendazole.^49^

A case study from 2017 revealed that 100 mg mebendazole taken twice daily for 5 consecutive days effectively terminated a T. trichiura infection.^50^

The timing of treatment in relation to meals can make a significant difference to treatment efficacy. The efficacy of albendazole (and possibly other anthelminthics) against hookworms can be enhanced by taking the drug on an empty stomach.^51^ When a single dose of 400 mg of albendazole was given to subjects who had not eaten for 6 hours or more prior to treatment, the cure rate for hookworm was raised from 59% to 90%.^52^ Different forms of tablet containing an anthelminthic drug may have different success rates. For example, chewable tablets may be more effective if they are crushed. ^53^

#### Mechanism of Action

As a vermicide, albendazole causes degenerative alterations in the intestinal cells of the worm by binding to the colchicine-sensitive site of β-tubulin, thus inhibiting its polymerization or assembly into microtubules (it binds much better to the β-tubulin of parasites than that of mammals). Albendazole leads to impaired uptake of glucose by the larval and adult stages of the susceptible parasites, and depletes their glycogen stores. Albendazole also prevents the formation of spindle fibers needed for cell division, which in turn blocks egg production and development; existing eggs are prevented from hatching.^54, 55^ Cell motility, maintenance of cell shape, and intracellular transport are also disrupted.^56^ At higher concentrations, it disrupts the helminths’ metabolic pathways by inhibiting metabolic enzymes such as malate dehydrogenase and fumarate reductase, with inhibition of the latter leading to less energy produced by the Krebs cycle. Due to diminished ATP production, the parasite is immobilized and eventually dies.^57, 58^

Some parasites have evolved to have some resistance to albendazole by having a different set of acids comprising β-tubulin, decreasing the binding affinity of albendazole.^59^

#### Pharmacokinetics

Oral absorption of albendazole varies among species, with 1–5% of the drug being successfully absorbed in humans, 20–30% in rats, and 50% in cattle.^60^

The absorption also largely depends on gastric pH. People have varying gastric pHs on empty stomachs, and thus absorption from one person to another can vary wildly when taken without food.^61^ Generally, the absorption in the GI tract is poor due to albendazole’s low solubility in water.^62^ Food stimulates gastric acid secretion, lowering the pH and making albendazole more soluble and thus more easily absorbed.^63^ Oral absorption is especially increased with a fatty meal, as albendazole dissolves better in lipids, allowing it to cross the lipid barrier created by the mucus surface of the GI tract. ^64^ To target intestinal parasites, albendazole is taken on an empty stomach in order to stay within the gut.^65^ Absorption is also affected by how much of the albendazole is degraded within the small intestine by metabolic enzymes in the villi.

Albendazole undergoes very fast 1st-pass metabolism in all species, such that the unchanged drug is undetectable in plasma. Most of it is oxidized into albendazole sulfoxide (also known as *ricobendazole* and *albendazole oxide*) in the liver by cytochrome P450 oxidases (CYPs) and a flavin-containing monooxygenase (FMO).^66, 67^ For systemic parasites, albendazole acts as a prodrug, while albendazole sulfoxide reaches systemic circulation and acts as the real antihelminthic. Albendazole sulfoxide is able to cross the blood-brain barrier and enter the cerebrospinal fluid at 43% of plasma concentrations; its ability to enter the central nervous system is what allows it to treat neurocysticercosis.

In humans, the metabolites mostly excreted in the bile, with only a small amount being excreted in the urine (less than 1%) and feces. The biological half-life of Albendazole and Mebendazole is 8-12 and 3-6 hours respectively.^68^ Like all benzimidazoles, albendazole has no residual effect, and thus does not protect well against re-infestations.

The drug used by the Department of Health in its deworming program is Albendazole. It is a drug that causes death of parasites by interfering with a protein found in parasites called tubulin. This would then inhibit the parasite from taking in glucose as its source for nutrients eventually leading to its death. The drug is active against tapeworms, roundworms, hookworms, threadworms, pinworms, pork worms, and whipworms.^69^

According to the World Health Organization, Albendazole (400 mg) is recommended as a public health intervention for all young children 12–23 months of age, preschool children 1–4 years of age, and school-age children 5–12 years of age. Preventive chemotherapy using this drug is intended to provide benefits only to infected individuals. However, uninfected individuals are treated also and this is only for logistical reasons. This way it has been proven to be the most cost–effective approach to reach infected individuals is to treat the entire group at risk without individual diagnosis.^70^

### Philippine Deworming Program Protocol

Preventive chemotherapy is one of the components of the helminth control program established by the World Health Organization (WHO).^71^ It is the process of administering regular anti-helminthic medications to individuals living in areas where soil-transmitted helminth (STH) infections are prevalent.^73, 74^ STH infections impair the physical and mental development of children which eventually leads to diminished educational advancement, reduced productive capacity and poverty.^74^ This tool is utilized to reduce the spread of these infections, eradicate the development of associated morbidity and improve the quality of life of individuals in impoverished areas.^31^ The purpose of repeated deworming treatment is to decrease the worm loads and prevent reinfection for successful elimination of the above-mentioned infections.^30^

Since school-age children are a high risk population for STH infections, their group is the main target of the deworming program. School intervention is considered a convenient and efficient method of mass deworming among school age-children. A single oral dose of either albendazole 400mg or mebendazole 500mg is given to each child. Praziquantel may also be given as an additional drug but dosage must be accurately determined according to the body weight or height of the child. ^1^For children below 24 months old, 1/2 dosage of albendazole is administered. Deworming program can be conducted annually or biannually. If baseline prevalence is greater than 50%, biannual administration is recommended._75_

The Department of Health in the Philippines conducts a National Deworming Month (NDM), a bi-annual mass drug administration for STH in public schools and communities every year during the months of January and July. The deworming program utilizes albedazole for all STH infections. The target population of the National School-Deworming Month (NDSM) are school-aged children from 5-18 years old, while the Community-Based Deworming Month (CBDM) targets pre-school children from 1-4 years old. STH infections are considered public health problems in the country since 66% of children ages 1-5 years old (2004, DOH-UP-UNICEF) and 54% of children ages 6-14 years old (2003, UP-CPH) were affected with STH.^76^ It is said that highest intensity of STH were found on children ages 1-2 years old, while children ages 5-12 years old were found to have the greatest load of infection.^77^ The main goal of the DOH NDM is to decrease STH prevalence in the country to less than 20% by year 2022.

In the Philippines, the Department of Health, in partnership with local government units and schools, give out anti-helminthic drugs during National Deworming Month, since STH has been a public health problem that has a damaging effect on the children’s health. These infections may lead to malnutrition, anemia, growth retardation, impaired cognitive functions, and other health problems, among school-aged children.^78^

In July 24 2015, the Department of Education made a memorandum (DM no. 80 s. 2015) containing the guidelines on the implementation of National Deworming Day, which is on the 29th of July. The program aims to deworm approximately 16 million school-aged children enrolled in all public schools in 1 day, to reduce the burden of STH infection with a positive impact on children’s education and health. 81 provinces in the country is said to be affected with STH.^77^ In the years 2004-2006, before implementation of the DM no. 80, National Deworming Program covered only 20% of affected school children, which is below the recommendation of WHO, which is 75%. Areas with the highest rates of infection were Leyte, wherein 70% of the adolescent population are infected with worms, Mindanao, and Samar. Infection rates in public schools are higher than in private schools, 60-70% and 10-20% respectivel.

### Conceptual Framework

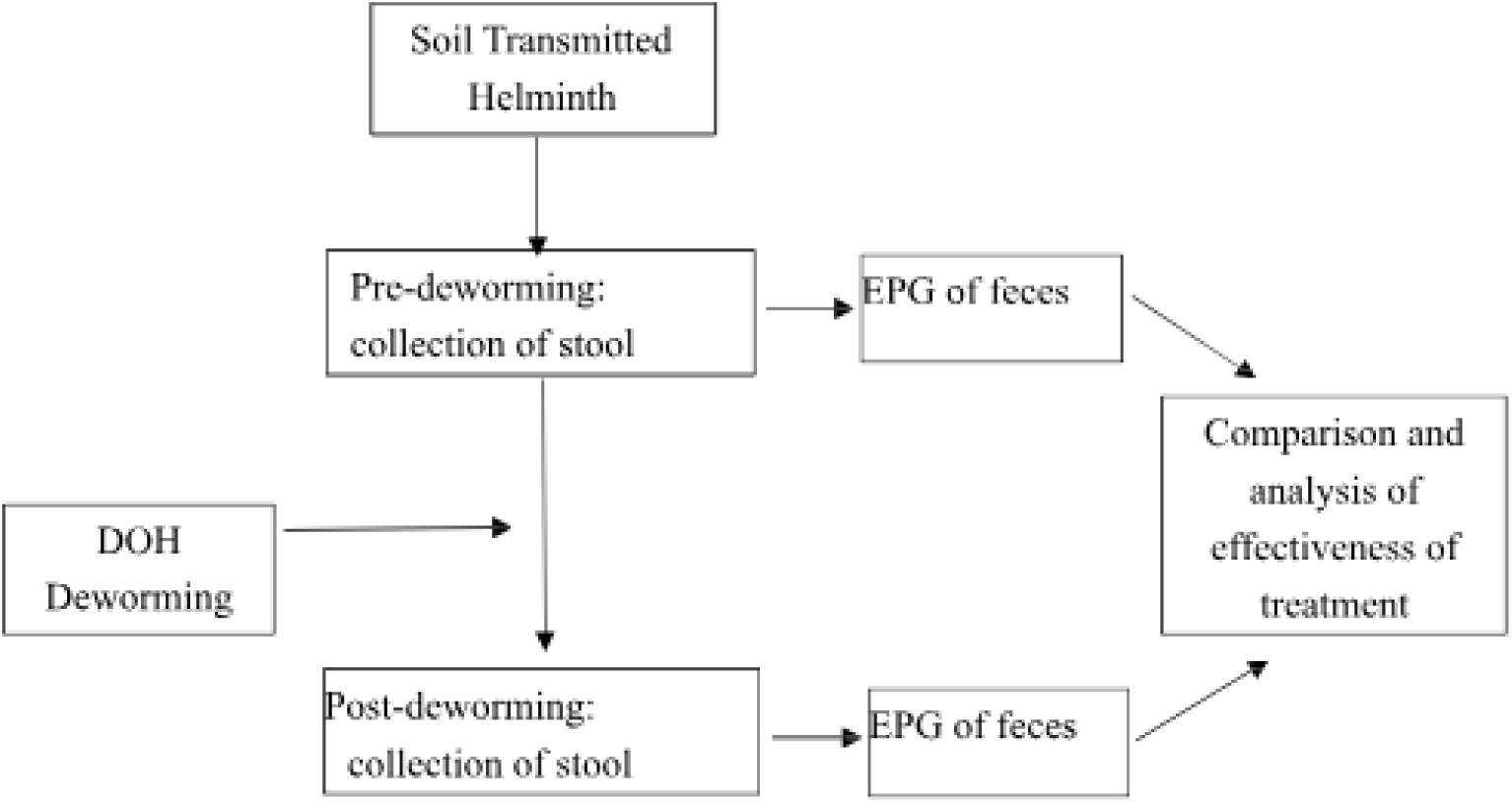

### Operational Definition of Terms

- Soil-transmitted helminths - comprises 3 types of nematodes or roundworms, specifically *Ascaris lumbricoides*, *Trichuris trichiura* and Hookworm species
- School-aged children - 9-12 year old students from 5^th^ grade who are enrolled in public schools under the DepEd-DOH deworming program
- Deworming program - bi-annual mass drug administration conducted during the months of January anxed July by the Department of Health among Philippine public schools
- Cure rate (CR) – proportion of subjects positive for parasites who become parasitologically negative after treatment. Refer to Eq [3]
- Eggs reduction rate (ERR) – refers to the reduction in the number of eggs excreted after treatment. Refer to Eq [3]

## Methodology

### Study Design

The present study is descriptive and comparative.

### Study Setting and Time Period

Collection of fecal samples from the students will be done in an Elementary School. Samples will be transported and analyzed in the Microbiology Laboratory of the Cebu Institute of Medicine. Duration of the study will span from July to September 2019.

### Study Population

#### Inclusion Criteria

All students aged nine to twelve years old from grade five of an Elementary school whose parents or guardians have consented to participate in the study and receive the deworming medications based on the DOH guidelines were included in this research study.

#### Exclusion Criteria

Unlabeled, contaminated and diarrheic stools were not examined. Respondents lost to follow-up were also excluded from the study.

#### Sample Size Calculation

We determined that the representative sample size of the study’s target population is equal to 188 respondents using Slovin’s formula of sampling size calculation. (Refer to Eq [1] in Appendix A)

### Study Maneuver

#### Sample Collection

In coordination with the Department of Health regarding the schedule for the Deworming program, permission to conduct the study was sought from the Superintendent of the Department of Education, as well as from the Principal of a certain public Elementary School. A letter of invitation was given to all Grade 5 students and their guardians for an orientation regarding the purpose of the study and the proper technique for specimen collection. Parents or guardians who were interested in enrolling their children in the study were asked to sign an informed consent and assent was also obtained from the participants. Specimen containers were then be distributed to those who agreed. Participating subjects were asked to provide a total of four stool specimens. Two of which were collected from July 16-26, 2019, and another two samples were collected fourteen to twenty-one days after the administration of the first cycle of deworming medications which was on July 29, 2019. The WHO recommends an interval of fourteen to twenty-one days between treatment and collection of follow-up data in order to ensure that eggs identified in a stool specimen are not from parasites that infected the subject after drug administration.^79^ Due to one reported death and hospitalization of 7 students from Surigao del Norte last July 31, 2019 (article link), the deworming program was suspended nationwide and resumed on September 4, 2019. As such, collection period from September 18-25, 2019 was set for students who were not able to participate in the July 29 deworming. Collected fecal samples were then transported to the Microbiology Laboratory at the Cebu Institute of Medicine for analysis.

#### Sample Processing

Prior to analysis, samples were confirmed on proper labelling with the subject’s name and assigned control number, and evaluated for the presence of contamination and stool consistency. Unlabeled, contaminated and diarrheic specimens were excluded from the study. The materials and reagents for processing of the samples were provided by the Department of Health. Analysis was done using the Kato-Katz method. The microscope slide was labeled with the subject’s control number and a template was placed on top of it. The fecal sample was homogenized before placing a small amount of fecal material on a piece of scrap paper. Filtration of the specimen was done by placing a nylon screen on top of the sample and scraping the upper surface using the flat sided portion of the spatula. The filtered feces was then added into the hole of the template and completely filling it. The template was removed carefully and the specimen was covered with a cellophane strip that was pre-soaked with methylene blue glycerol solution. Even distribution of fecal material was achieved by pressing a clean microscope slide against the cellophane covered area of the sample slide and gently sliding it sideways. Slides were examined in a systematic manner, making sure that all fields were read. For optimal identification of Hookworm eggs, slides were read within thirty minutes to one hour, whereas ova of *Ascaris lumbricoides* and *Trichuris thrichiura* may remain recognizable for up to several months. As part of the quality control process, a reference microscopist was present to verify positive findings. 10% of the slides was blindly re-examined.^80^ The results of this study will be reported to the following: Dean of the College of Medicine in CIM, chosen Elementary School, Department of Education and Department of Health for appropriate intervention, if necessary.

## Plan for Analysis

### Statistical Analysis

The descriptive aspect of the study design aims to describe the percentage and intensity of STH infection among the respondents. As such, Helminths will be identified by species, quantitated as eggs per gram of stool and recorded in the data collection forms (refer to Appendix B).

Percentage of STH infection will be determined using Eq [2]

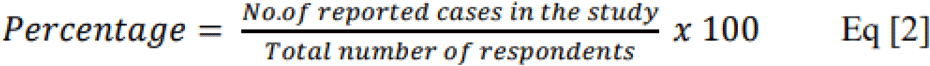

Intensity of the STH infection was categorized into light, moderate and heavy intensity according to species-specific threshold values set by the World Health Organization (See Appendix A).

McNemar’s test was used to determine if there is a significant difference in terms of percentage of STH infection before and after the deworming program using SPSS version 26. A p-value of 0.05 was considered the limit of statistical significance.

To further determine the effectiveness of Albendazole, the total Cure rate (CR) and Eggs reduction rate (ERR) along with the CR and ERR for each species of soil transmitted helminths found in the fecal samples were calculated using Eq [3]

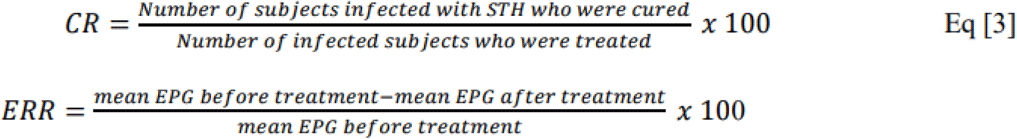

### Effectiveness of the treatment is: ^80^

- satisfactory if CR is >90% and ERR is greater than or equal to the reference value (Appendix A)
- doubtful if the ERR is less than or equal to the reference value by <10%
- reduced if the ERR is less than the reference value by at least 10%

## Results and Discussion

A total of 92 fecal samples were collected at the Public Elementary School and were examined in the Cebu Institute of Medicine microbiology laboratory from July 2019 to September 2019. Out of the 356 grade 5 students who were invited to participate in the study, a total of 53 respondents submitted fecal samples for baseline determination of STH infection. The 4 respondents who were lost to follow-up were excluded from the study. From these subjects, 6 (12.24%) were 9 year-olds, 39 (79.59%) were 10 year-olds, 3 (6.12%) were 11 year-olds, and 1 (2.04%) was a 12 year-old. (Fig. 1). There were 26 (53.06%) males, and 23 (46.94%) females. (Fig. 2)

**Figure 1.**
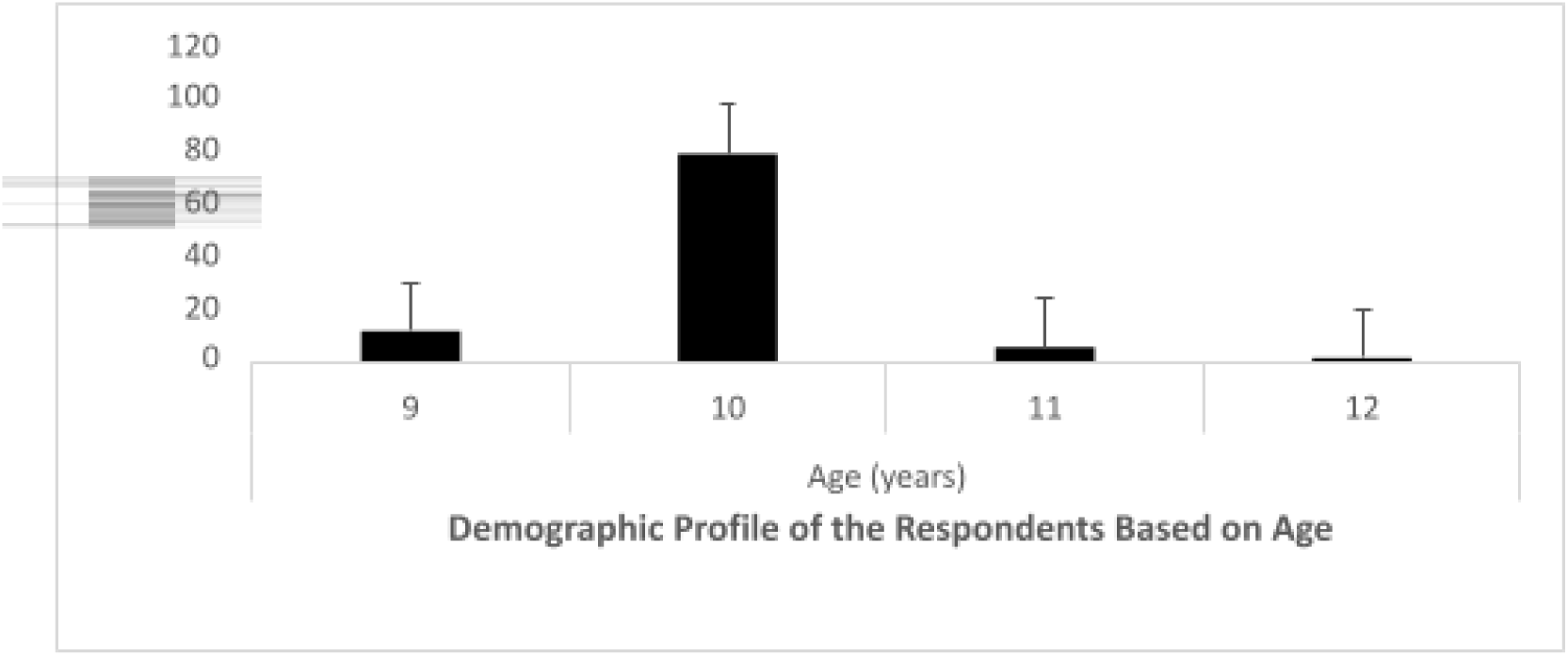
Demographic Profile of the Respondents based on Age.

**Figure 2.**
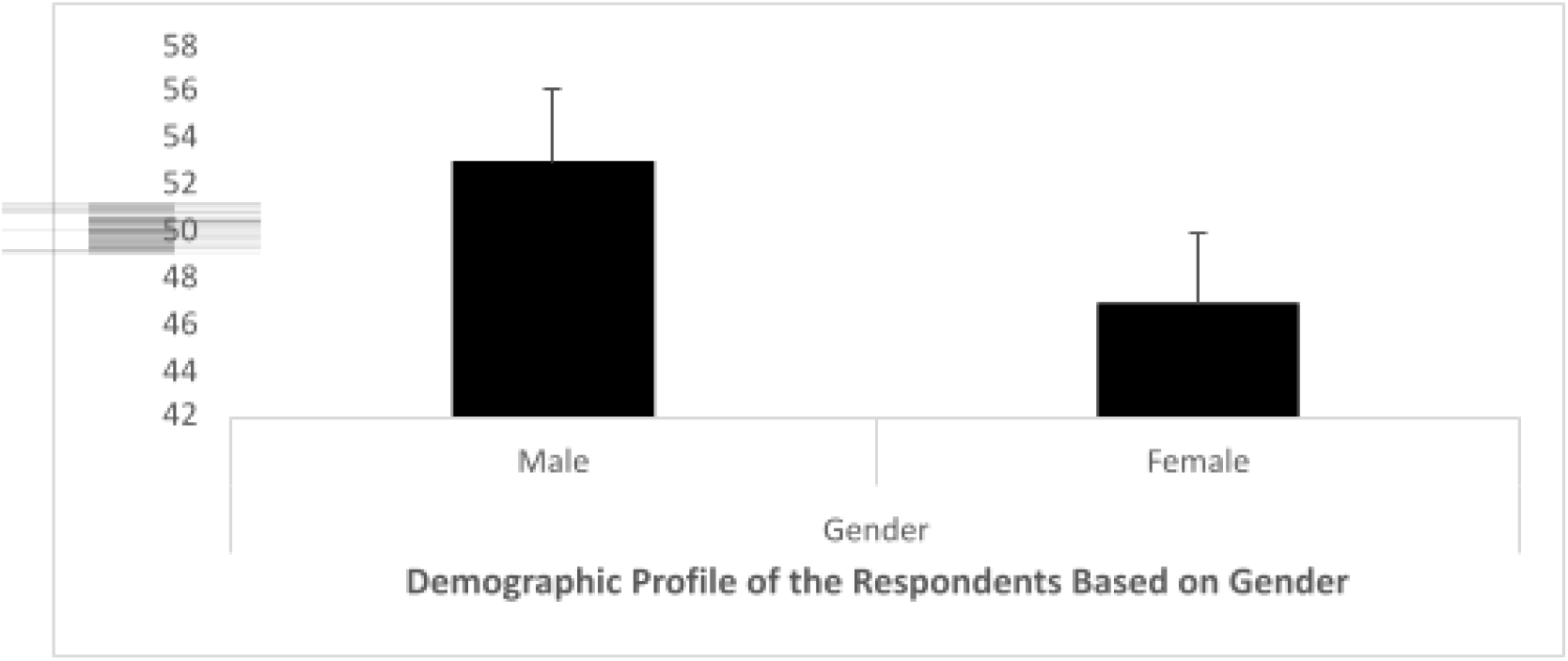
Demographic Profile of the Respondents based on Gender.

The most abundant age group among the respondents were 10 year-olds. Soil-transmitted helminth (STH) infections are the most common parasitic infections among school-children worldwide, especially among impoverished communities. In the Philippines, the prevalence in school-aged children with STH reportedly was as high as 67% in 2001.

A number of 188 participants were required to participate in the study, as computed using Slovin’s formula (refer to appendix A). However, only 53 participated and 49 were included in the study. This is primarily due to lack of consent from the parents of the students. Furthermore, the alleged recent deaths due to the deworming program^82^ and Dengvaxia^83^ in the Philippines have contributed to the negative perspective of the general population towards DOH programs, and thus remained as a major concern for the parents who then opted not to participate in the deworming activity.

Out of the 49 participants, a total of 10 (20.41%) respondents were positive for soil transmitted helminths. The decline in the prevalence of STH infection may be attributed to history of past chemotherapy since the bi-annual National school-based deworming program of the Department of Health involving all public elementary schools is now in its fourth year of implementation. Out of the 49 participants, a total of 10 (20.41%) respondents were positive for soil transmitted helminths. The decline in the prevalence of STH infection may be attributed to history of past chemotherapy since the bi-annual National school-based deworming program of the Department of Health involving all public elementary schools is now in its fourth year of implementation.

Among the soil transmitted helminths, only the ovas of *Ascaris lumbricoides* and *Trichuris trichiura* were observed under the light microscope (Fig. 3). Both fertilized and unfertilized *Ascaris lumbricoides* eggs are passed in the stool of the infected host. Fertilized eggs are rounded and have a thick shell with an external mammillated layer that is often stained brown by bile. In some cases, the outer layer is absent (known as decorticated eggs). Fertile eggs range from 45 to 75 µm in length. Unfertilized eggs are elongated and larger than fertile eggs (up to 90 µm in length). Their shell is thinner and their mammillated layer is more variable, either with large protuberances or practically none. Unfertilized eggs contain mainly a mass of refractile granules.^84^ Meanwhile, Trichuris trichiura eggs are 50-55 micrometers by 20-25 micrometers. They are barrel-shaped, thick-shelled and possess a pair of polar “plugs” at each end. And are unembryonated when passed in the stool.^85^ No Hookworm spp. ova were found. The observation of A. lumbricoides and T. trichiura in the sample population can be linked to the high endemicity and widespread distribution of both species as compared to hookworm infections that are more circumscribed to a smaller foci in Visayas and Mindanao.^86^

**Fig. 3.**
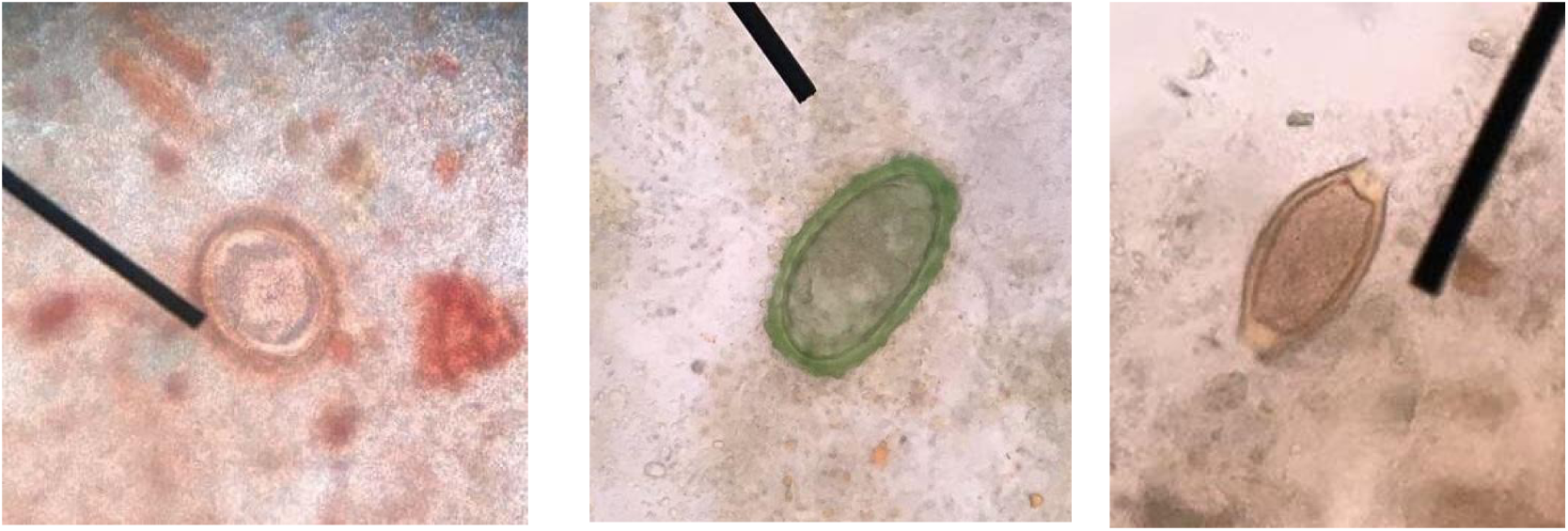
A*s*caris eggs Fertilized (A) and Unfertilized (B), *Trichuris trichiura* (C) under light microscope.

Sensitivity estimates of the Kato-Katz technique for one stool sample were 96.9%, 65.2% and 91.4%, for *A. lumbricoides*, hookworm and *T. trichiura*, respectively. Specificity estimates for one stool sample were 96.1%, 93.8% and 94.4%, for *A. lumbricoides*, hookworm and *T. trichiura*, respectively. Low sensitivity of the Kato-Katz for detection of hookworm infection may be related to rapid degeneration of delicate hookworm eggs with time.^87^ However, there were incidental findings of *Enterobius vermicularis* in 3 respondents. (Fig. 4). Adhesive tape test remains so far the method of choice for diagnosing *E. vermicularis*.^88^ This is due to the specific egg-shedding habits of *E. vermicularis* worms, in which the eggs would be found in the first portion of stool passed after the night. The eggs of *E. vermicularis* are transparent, elongate to oval in shape, and slightly flattened on one side. They measure 50—60 µm by 20—30 µm and are usually partially embryonated when shed. The principal mode of transmission of *E. vermicularis* is self-inoculation, also called Autoinfection, which happens when the hands that scratched perianal area comes in contact with the mouth or through exposure to eggs in the environment like contaminated surfaces, clothes, bed linens, etc. For this reason, it is prevalent in both temperate and tropical regions both in developed and underdeveloped countries infecting 208.8M worldwide. The people most likely to be infected are children under 18, people who take care of infected children and people who are institutionalized.^89^ Local prevalence of 29% from exclusive private schools, 56% from public schools. Higher in females (16%) compared to males (9%).

**Figure 4.**
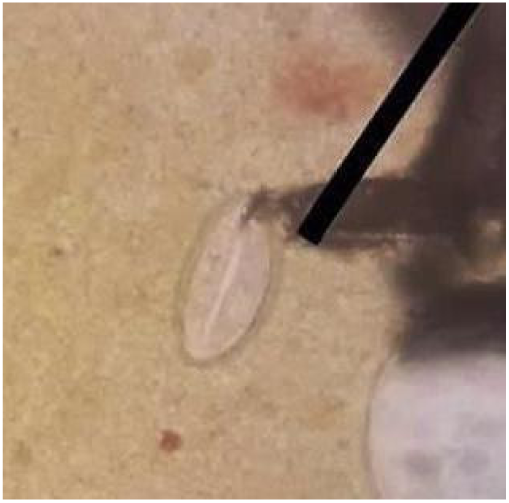
*Enterobius vermicularis* under light microscope.

Among the types of soil transmitted helminths found at baseline, *A. lumbricoides* was the most frequent species isolated (18.37%) followed by *T. trichiura* (6.12%) (Table A1.3, A1.4). Single infection with *A. lumbricoides* (12.24%) was the most prevalent among the respondents, followed by co-infection with *A. lumbricoides* and *T. trichiura* (4.08%) and single infection with *T. trichiura* (2.04%) was found to be the least prevalent. were also incidental findings of single infection with E. vermicularis (4.08%) and co-infection of *E. vermicularis* with *A. lumbricoides* (2.04%) (Fig. 5).

**Figure 5.**
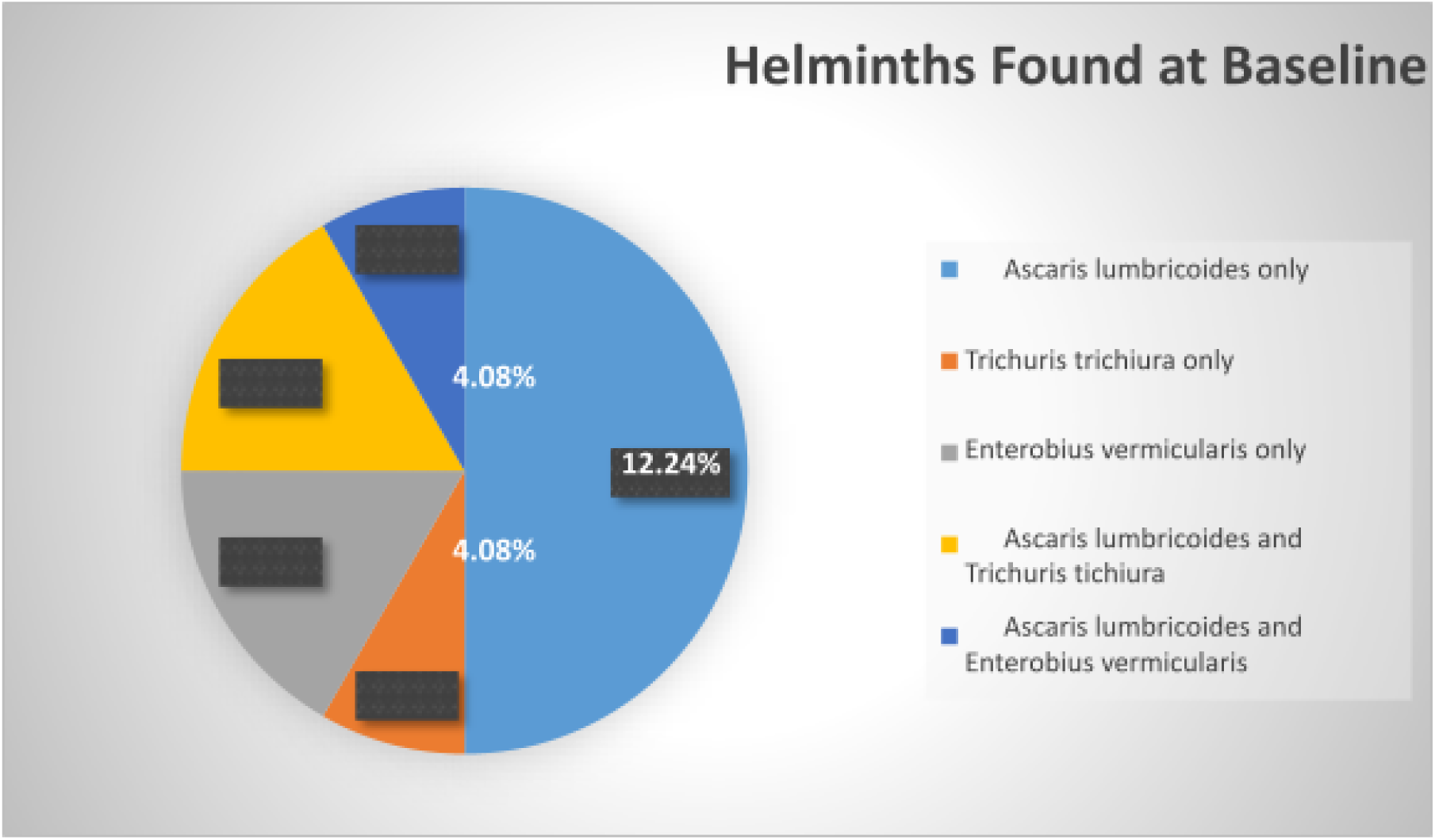
Helminths Found at Baseline.

The Department of Health Research Institute of Tropical Medicine conducted a study last February 2017 to determine the prevalence of soil-transmitted helminth infections using Kato-Katz technique among children 5-16 years old. The results of the study are shown below:

**Table 1.**
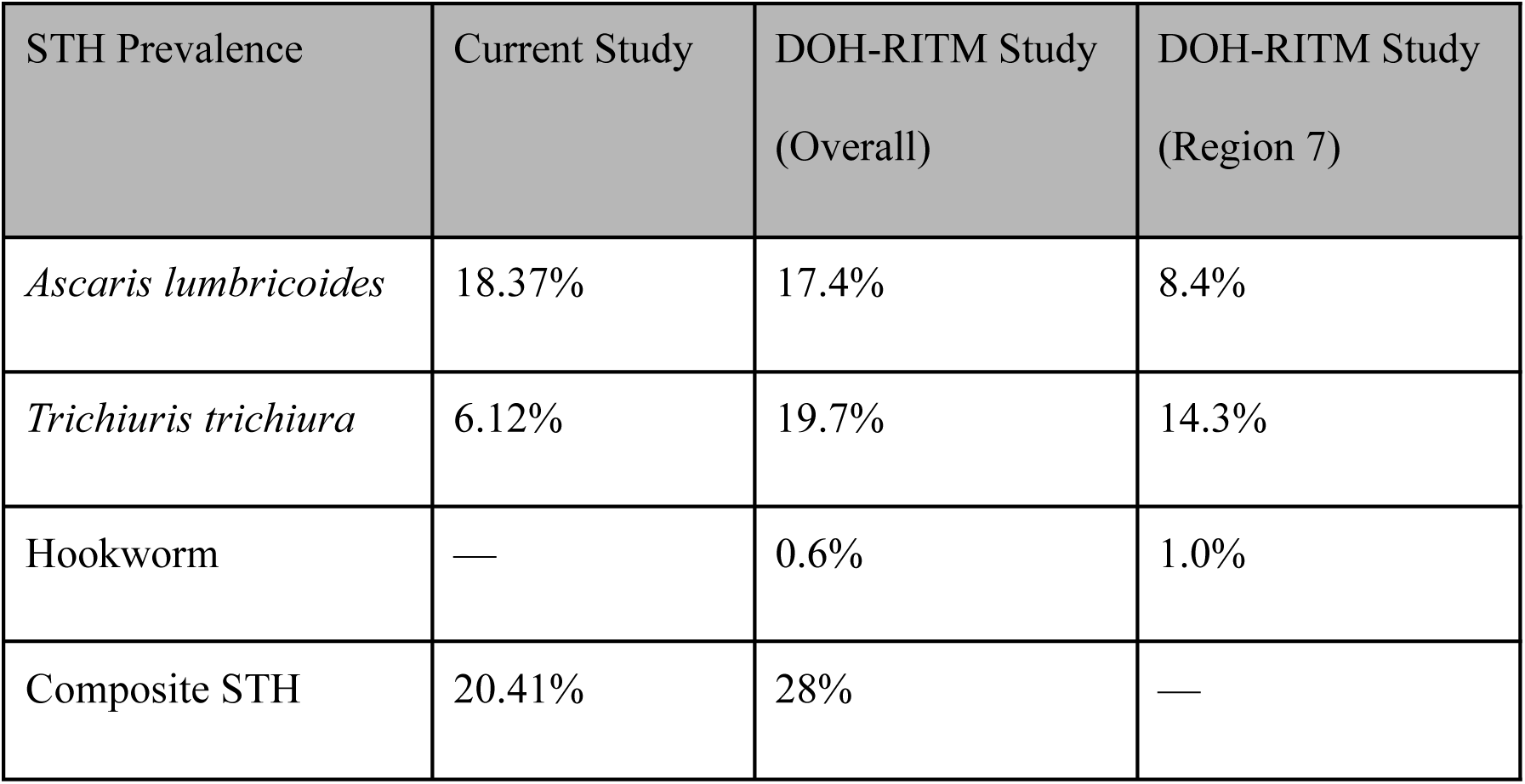
A Comparison of results in the current study and the DOH-RITM study conducted last February 2017.

In the study conducted by DOH RITM, *Ascaris lumbricoides* infection was found to be highest in the age group 5-7 years old with 19% and lowest in the age group 10-16 years old with 16.5%. *Trichiuris trichiura* infection, on the other hand, was found to be highest in the age group 10-16 years old with 21.1% and lowest in the age group 5-7 years old with 17%. The variation in the results of both studies may be due to the current study’s low student participation, the age of the participants, and the number of schools participating.

Stratification of STH intensity by WHO standards is according to the number of helminth eggs per gram of excreted human feces (Table A1.2). Data regarding the percentage of light, moderate and heavy intensity infection before and after treatment are summarized in Table 2. Majority of the infections before and after treatment were of light intensity at 16.33% and 4.08%, respectively. Only 4.08% were classified under moderate intensity, and none were found with heavy intensity infection. Specifically, light intensity Ascaris infection was found to be the highest (14.29%) at baseline whereas light intensity Trichuris infection (4.08%) was more prevalent during follow-up. (Fig. 6)

**Table 2.** Intensity of STH Infection.

| Parameters (n=49) | Light Intensity |  | Moderate Intensity |  | Heavy Intensity |  |
| --- | --- | --- | --- | --- | --- | --- |
|  | f | % | f | % | f | % |
| **Composite STH infection** |  |  |  |  |  |  |
| Before treatment | 8 | 16.33 | 2 | 4.08 | 0 | 0 |
| After treatment | 2 | 4.08 | 0 | 0 | 0 | 0 |
| ***Ascaris lumbricoides*** |  |  |  |  |  |  |
| Before treatment | 7 | 14.29 | 2 | 4.08 | 0 | 0 |
| After treatment | 0 | 0 | 0 | 0 | 0 | 0 |
| ***Trichuris trichiura*** |  |  |  |  |  |  |
| Before treatment | 3 | 6.12 | 0 | 0 | 0 | 0 |
| After treatment | 2 | 4.08 | 0 | 0 | 0 | 0 |

**Figure 6.**
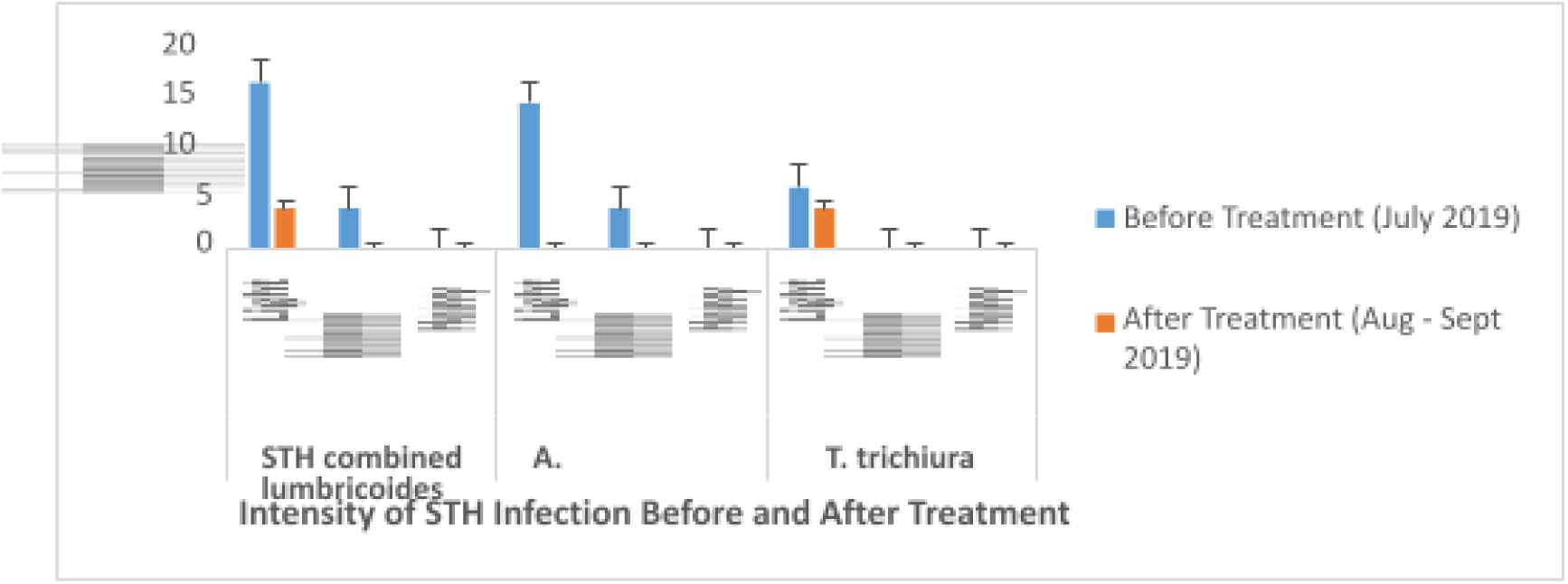
Intensity of STH Infection Before and After Treatment.

The severity of the clinical manifestations of helminth infections are positively correlated with the intensity of infection. Most individuals with Ascaris infection are often asymptomatic especially those with light intensity infections. For those with moderate intensity and heavy intensity infections, either pulmonary or intestinal manifestations are seen. Pulmonary ascariasis also known as Loeffler syndrome commonly occurs in those without prior ascaris infection but is rare in highly endemic areas. It may manifest as dry cough, dyspnea, fever, wheezing, substernal discomfort, and blood-tinged sputum. These pulmonary manifestations are observed during the early phase of the infection (four to sixteen days after ingestion of infective eggs), and are usually self-limited. On the other hand, intestinal manifestations are observed during the late phase of the infection (six to eight weeks after ingestion of infective eggs). Moderate intensity infections may manifest as abdominal discomfort, anorexia, nausea, vomiting, and diarrhea. Nutrient losses associated with nitrogen, albumin and other proteins, fat, lactose and Vitamin A have also been reported. Heavy intensity infections may manifest as severe abdominal pain, fatigue, vomiting, and weight loss. Vomiting and passage of macroscopic adult worms and complications such as intestinal obstruction, volvulus, ileocecal intussusception, gangrene, intestinal perforation, malnutrition, hepatobiliary and pancreatic involvement may occur in those with heavy intensity infections.^90^

Similar to ascaris infection, most individuals with light intensity Trichuris infection are asymptomatic. Some may experience right lower quadrant or vague periumbilical pain. When associated with ascaris or hookworm infection, symptoms such as epigastric pain, vomiting, distention, flatulence, anorexia and weight loss may occur. Moderate infections can result in chronic trichuris colitis, while heavy infection can cause Trichuris dysentery syndrome (TDS), massive infantile trichuriasis and iron deficiency anemia (IDA). Manifestations of TDS include abdominal distention, diarrhea and severe dysentery, pallor and anemia, growth impairment, and rectal prolapse. Massive infantile trichuriasis typically occurs in children aged 3 to 10 years old and may manifest as hypoproteinemia, severe anemia and clubbing of fingers. Chronic heavy infections that occur during childhood have detrimental effects on cognition. The estimated mean blood loss per day per *T. trichiura* is 0.005 mL and may result in iron deficiency anemia which is often aggravated by hookworm coinfection.^91, 92^

During follow-up determination, 2 respondents (4.08%) were found to be still infected with *T. trichiura*, obtaining an overall reduction of 16.33% from baseline after treatment with a single dose of 400 mg Albendazole. (Fig. 7)

**Figure 7.**
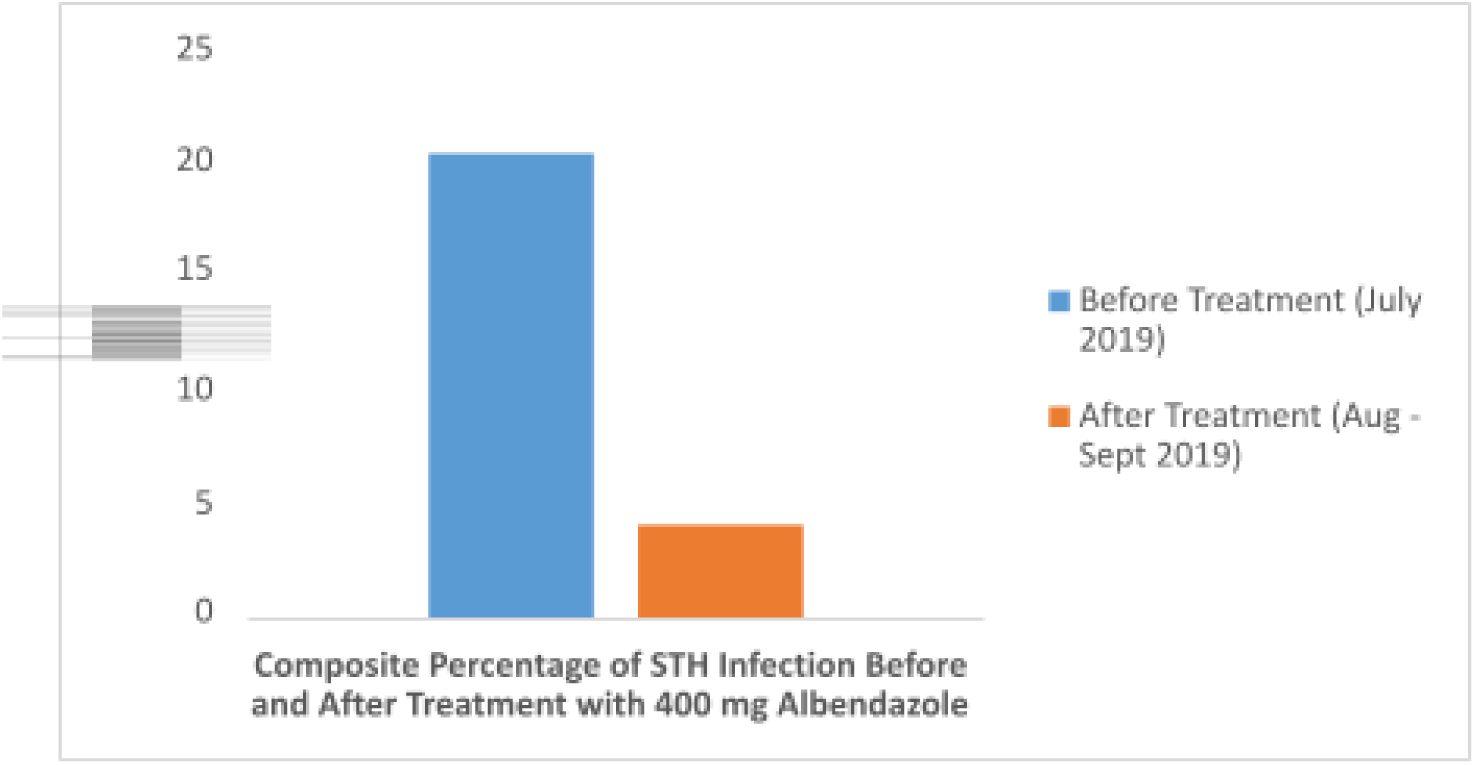
Composite Percentage of STH Infection Before and After Treatment.

There is a significant decline (p=0.008) in the overall percentage of soil-transmitted helminth infection among the respondents. These findings are consistent with a previous study done in Cavite province, Philippines, wherein a significant (22.3%) reduction in overall STH prevalence was obtained during the follow-up assessment.^18^ Our results also show that 14-21 days after treatment, the percentage of *A. lumbricoides* decreased significantly (McNemar test, p<0.001), while there was no significant decrease in the percentage of *T. Trichiura* (McNemar test, p=1.000) (Fig. 8). These support the findings of two other studies in Indonesia where post-treatment reduction of *Ascaris lumbricoides* was significant; however, both studies also showed that single dose Albendazole was unable to reduce the prevalence of *Trichuris trichiura* significantly.^93, 94^

**Figure 8.**
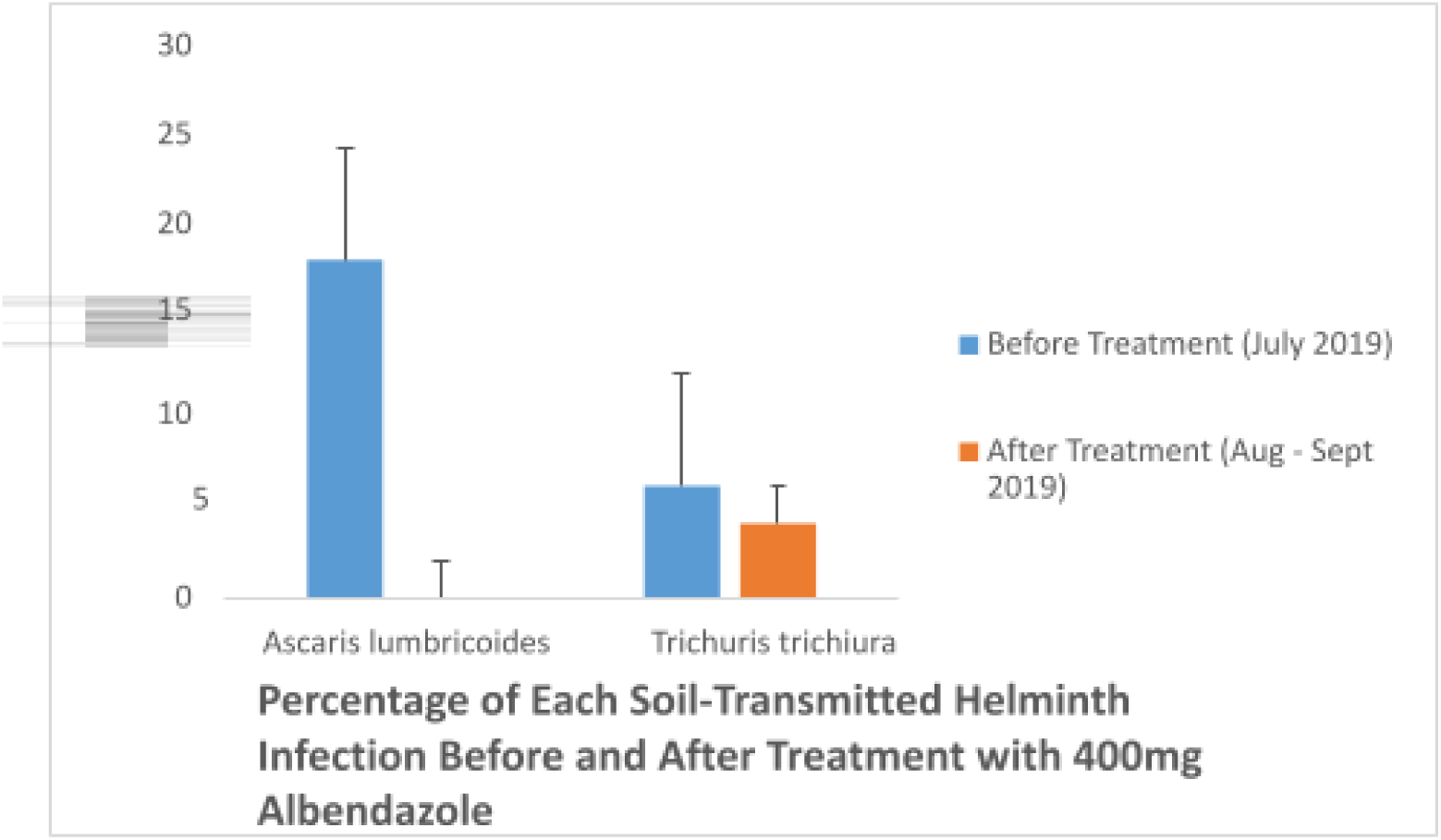
Percentage of Each Soil-Transmitted Helminth Infection Before and After treatment.

Furthermore, despite the decrease of *Ascaris* infection among the respondents, *Trichuris* infection was still present even after treatment. According to CDC, treatment with anthelmintic therapy, like Albendazole, is found to be effective on Trichuriasis when taken for 3 days, 400 mg dosage.^95^ Table 3 shows that the effect of single-dose 400mg Albendazole on Trichuriasis in terms of egg reduction rate (ERR) is 51.02% whereas only a cure rate (CR) of 33.33% was achieved. This means that the drug is effective in reducing the viable eggs in an infected patient but this dosage is not sufficient to cure patients infected with *Trichuris trichiura*.

**Table 3.** Cure Rate and Egg Reduction Rates.

| Variable | Helminths |  |  |
| --- | --- | --- | --- |
|  | *Ascaris lumbricoides* | *Trichuris trichiura* | Composite STH Infection |
| Number of positive responden |  |  |  |
| Before treatment | 9 | 3 | 10 |
| After treatment | 0 | 2 | 2 |
| **Cure Rate (%)** | 100 | 33.33 | 80 |
| Arithmetic Mean EPG |  |  |  |
| Before treatment | 78.67 | 196 | 108 |
| After treatment | 0 | 96 | 96 |
| **Egg Reduction Rate (%)** | 100 | 51.02 | 11.11 |

A study from Iran was conducted regarding the efficacy of albendazole and mebendazole on STH infections. The results of these studies also support the unsatisfactory use of single dose Albendazole on patients with trichuriasis.^96^ Another study in Cape, South Africa showed a cure rate of 23% with a 400mg dose of Albendazole, 56% with an 800mg dose of Albendazole, and 67% with 1200mg dosage. Egg reduction rates were 96.8%, 99.3% and 99.7% for the respective dosages mentioned.^97^ Besides the actual effect of the drug to the helminths, other factors could also affect the prevalence of these STH infections on different populations. Intensity of infection on each individual could affect treatment of current signs and symptoms. People with heavier infections may need surgical treatment as an option. Distribution of medicinal treatments also depends on the prevalence of the infections in the area. Prevalence of >50% would mean that treatments should be done twice a year in order to reduce infection.^98^

## Limitations

The study employs less number of participants than what is statistically required. The factors that led to parents not participating in the study include the altered perspective of the public towards the deworming program as a consequence of recent death of a nine-year old pupil in Gigaquit, Surigao del Norte allegedly after taking the deworming drug on July 23, 2019 and many preceding events involving deaths of young children that resulted from Dengvaxia administration in 2018. The DOH has reported difficulties in successfully conducting the deworming programs for the same reason as the parents are unwilling to let their children be dewormed. Furthermore, collaborating with a field expert and a more subtle approach to persuade the parents to allow their children to participate in the study could have resulted in the achieving the required number of participants for the study.

## Conclusion

A single dose of Albendazole 400mg was effective against Ascariasis with both the egg reduction rate and cure rate yielding 100%. The drug was also effective against Trichuriasis based on its egg reduction rate of 51.02%; however, it was less effective or insufficient based on its cure rate of 33.33%.

## Recommendations

The researchers recommend that the DOH should re-evaluate their protocol to address the Trichuris trichiura infection and prevent its reinfection. This can be achieved by increasing the duration of the treatment from single dose to triple dose. Since there is no equal distribution of STH infections across the country, a spatially targeted approach in STH intervention can also be employed to specifically target areas with higher prevalence and intensity of infection. Follow-up surveillance by the local government units must also be done to assess and monitor the effectiveness of the deworming program. It is also recommended that proper education towards stakeholders be emphasized so as to increase awareness and participation of students in the National Deworming Program and future studies. With higher participation rates, prevalence of STH in the Philippines will be more accurate and there will be better monitoring of the effectiveness of the Deworming Program. For future studies, the researchers recommend that questionnaires be included to address the possible roles of habits, environmental factors, and other possible risk factors in the causation of STH infection. Improvements in sanitation, water supply and hygiene within the household and in the community should also be made in order to interrupt the parasite life cycle and achieve sustainable control of infection. Lastly, since incidental findings of *Enterobius vermicularis* ova were found, we recommend future researchers who want to focus on this parasite to employ the scotch tape method for specimen collection for better isolation and correct quantitation of the prevalence of this parasite.

## Data Availability

All data produced in the present work are contained in the manuscript

## Appendix A Slovin’s formula for sample size calculation

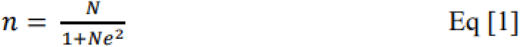

Where: n = sample size

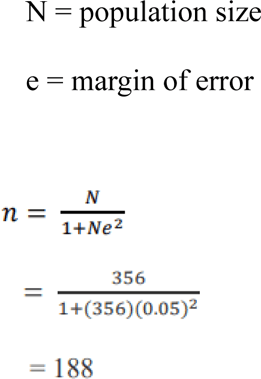

**Table A1.1.** Thresholds for the classification of intensity of STH infections in individuals.

| Organism | Light intensity | Moderate intensity | Heavy intensity |
| --- | --- | --- | --- |
| *Ascaris lumbricoides* | 1 – 4,999 eggs per gram | 5,000 – 49 999 eggs per gram | >/ 50,000 epɡ |
| *Trichuris trichiura* | 1 – 999 epɡ | 1,000 – 9,999 epɡ | >/ 10,000 epɡ |
| **Hookworm** | 1 – 1,999 epɡ | 2,000 – 3,999 epɡ | >/ 40,000 epɡ |

(Source: WHO, 1998)

**Table A1.2.** Reference drug efficacy of albendazole (chewable tablet, 400 mg)

| Organism | Reference efficacy (egg reduction rate, %) |
| --- | --- |
| *Ascaris lumbricoides* | ≥ 95 |
| **Hookworm** | ≥ 90 |
| *Trichuris trichiura* | ≥ 50 |

(Source: WHO, 2013)

**Table A1.3.** Intensity of Ascariasis According to the Demographic Profile of the Respondents Before Treatment.

(Source: WHO, 2013)
|  |  |  |  |  |  |  |  |
| --- | --- | --- | --- | --- | --- | --- | --- |
| **Variables** | **Classification** | Light Intensity (1-4,999 EPG) | | Moderate Intensity (5, 000-49,999 EPG) | | Heavy Intensity ($\geq 50,000$ EPG) | |
|  |  | f | % | f | % | f | % |
| **Age** | 9 year-old | 0 | 0 | 0 | 0 | 0 | 0 |
|  | 10 year-old | 4 | 8.16 | 2 | 4.08 | 0 | 0 |
|  | 11 year-old | 2 | 4.08 | 0 | 0 | 0 | 0 |
|  | 12 year-old | 1 | 2.04 | 0 | 0 | 0 | 0 |
| **TOTAL** |  | 7 | 14.29 | 2 | 4.08 | 0 | 0 |
| **Gender** | Female | 2 | 4.08 | 1 | 2.04 | 0 | 0 |
|  | Male | 5 | 10.20 | 1 | 2.04 | 0 | 0 |
| **TOTAL** |  | 7 | 14.29 | 2 | 4.08 | 0 | 0 |

**Table A1.4.** Intensity of Trichuriasis According to the Demographic Profile of the Respondents Before Treatment.

|  |  |  |  |  |  |  |  |
| --- | --- | --- | --- | --- | --- | --- | --- |
| **Variables** | **Classification** | Light Intensity (1-999 EPG) | | Moderate Intensity (1,000-9,999 EPG) | | Heavy Intensity ($\geq 10,000$ EPG) | |
|  |  | f | % | f | % | f | % |
| **Age** | 9 year--old | 0 | 0 | 0 | 0 | 0 | 0 |
|  | 10 year—old | 2 | 4.08 | 0 | 0 | 0 | 0 |
|  | 11 year—old | 0 | 0 | 0 | 0 | 0 | 0 |
|  | 12 year—old | 1 | 2.04 | 0 | 0 | 0 | 0 |
| **TOTAL** |  | 3 | 6.12 | 0 | 0 | 0 | 0 |
| **Gender** | Female | 2 | 4.08 | 0 | 0 | 0 | 0 |
|  | Male | 1 | 2.04 | 0 | 0 | 0 | 0 |
| **TOTAL** |  | 3 | 6.12 | 0 | 0 | 0 | 0 |

## Appendix B Data Collection Form

***Note:*** *The sample/patient IDs used in this study were not known to anyone outside the research group, and therefore do not pose a risk of identifying study subjects*

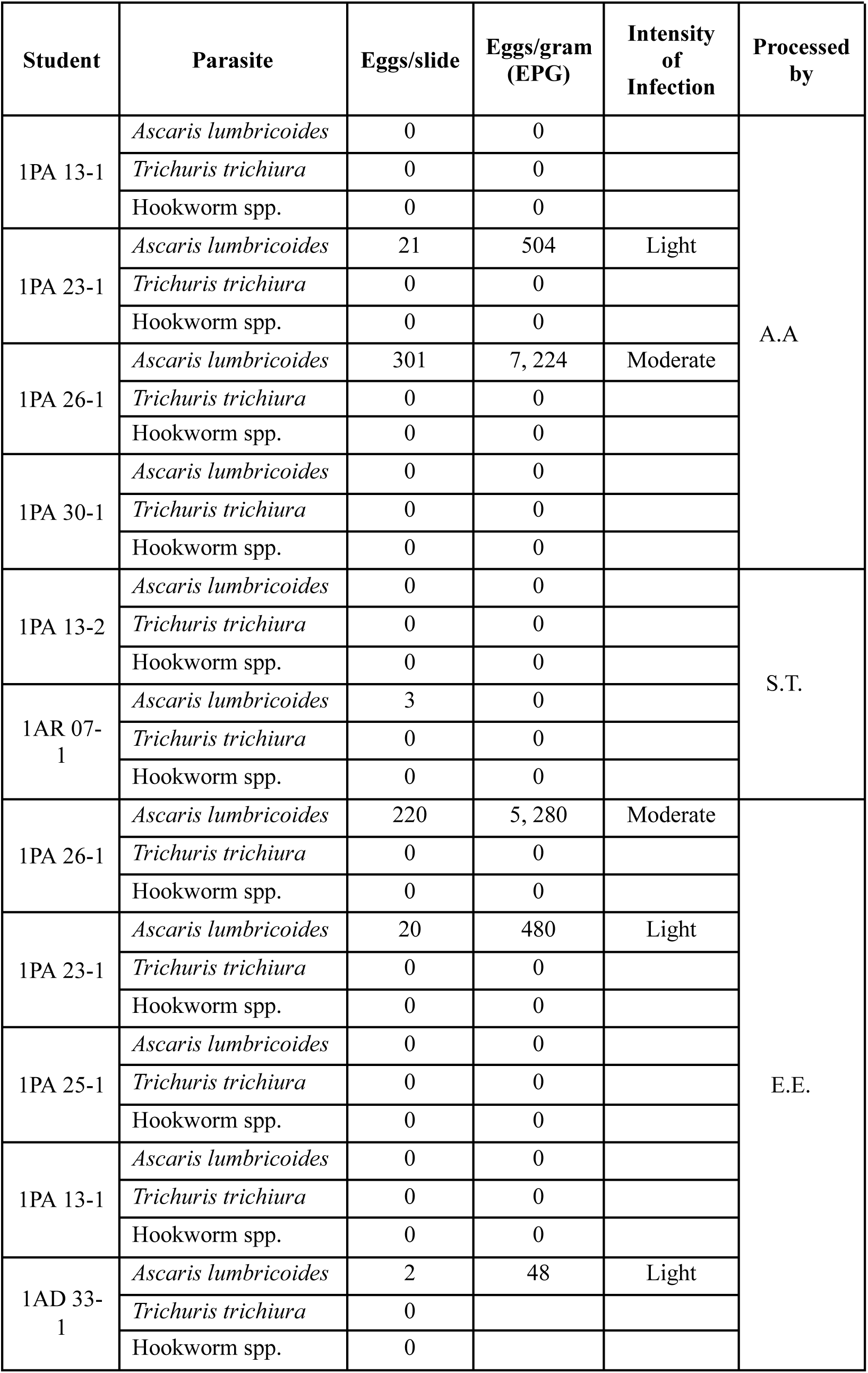
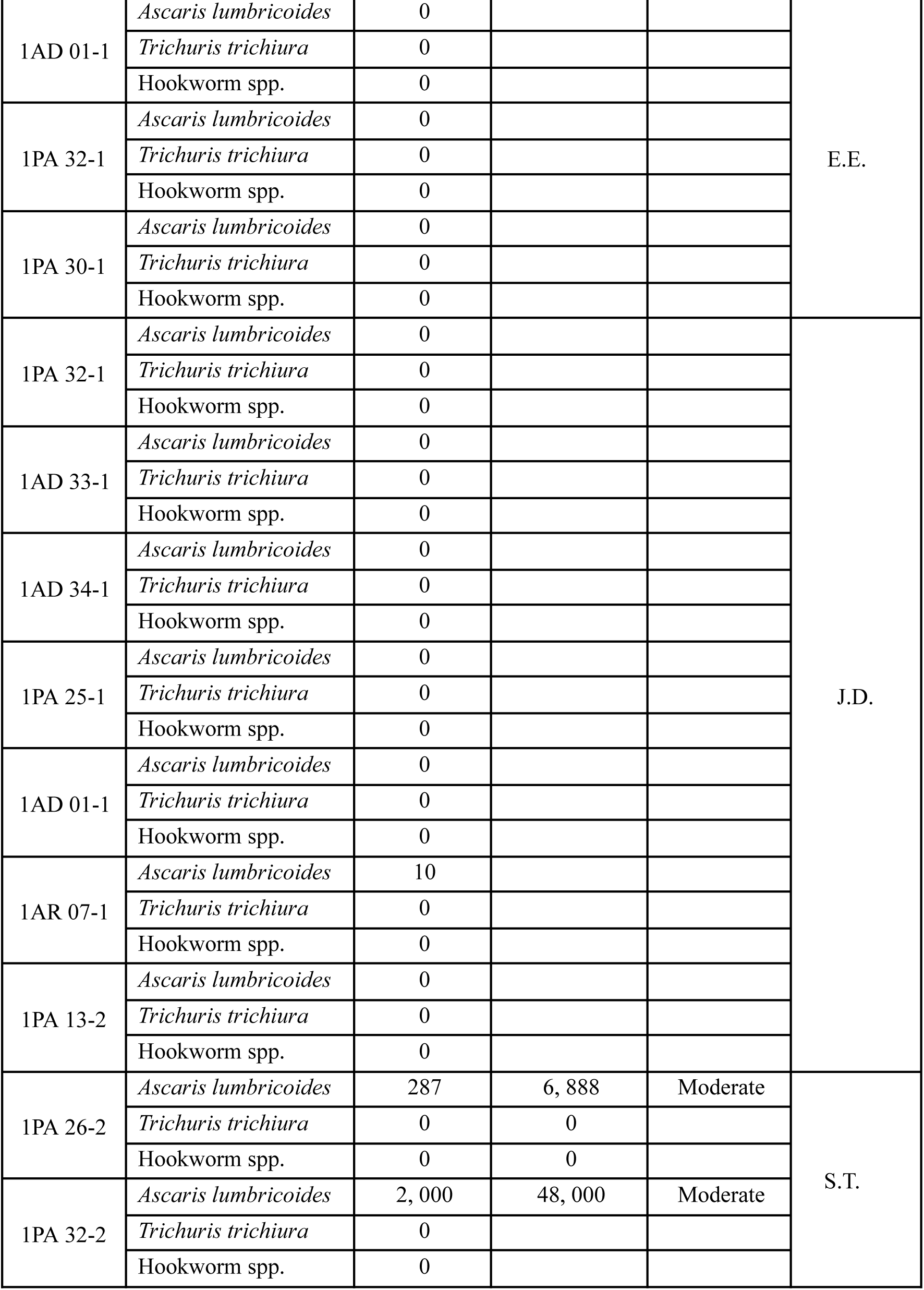
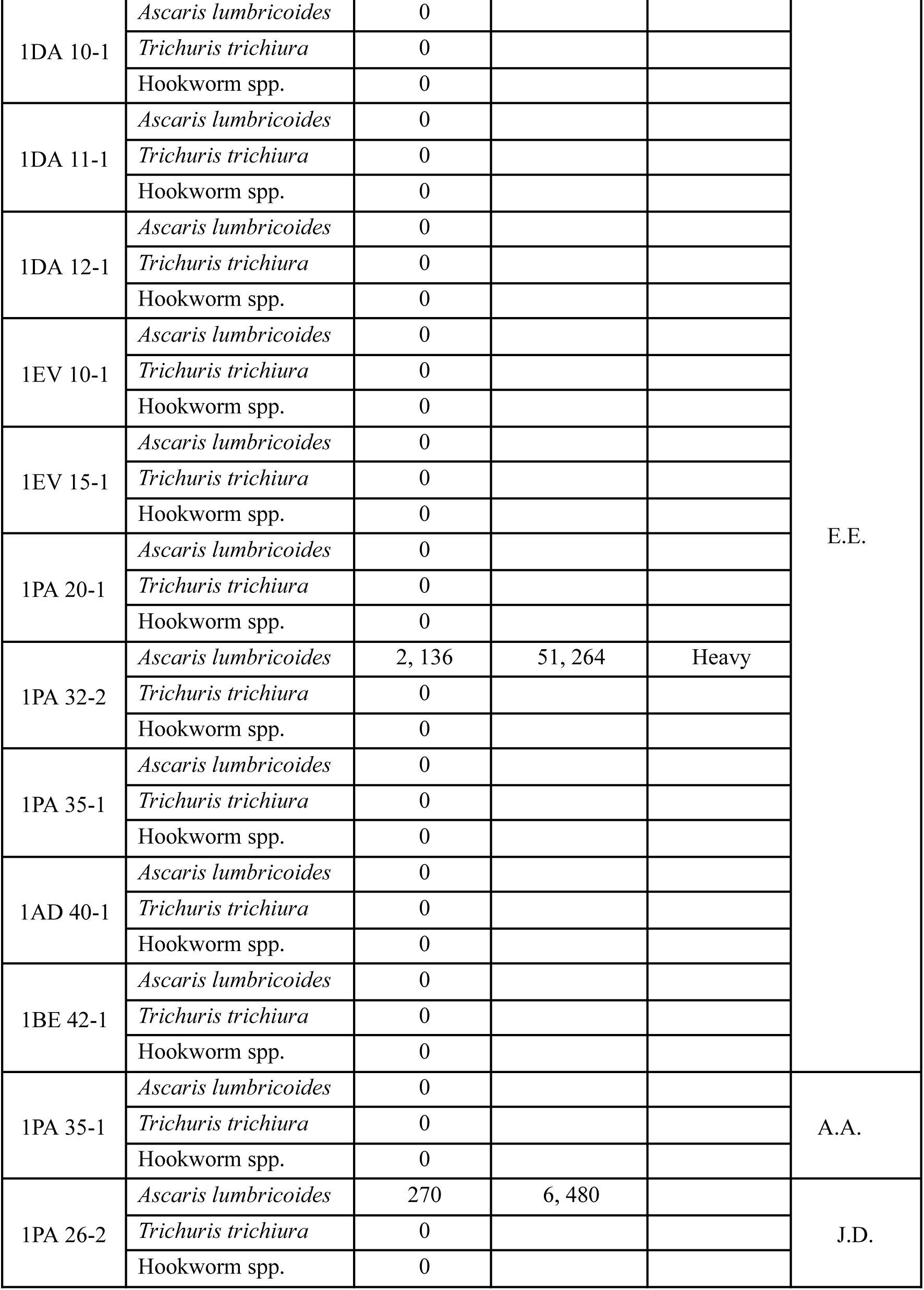
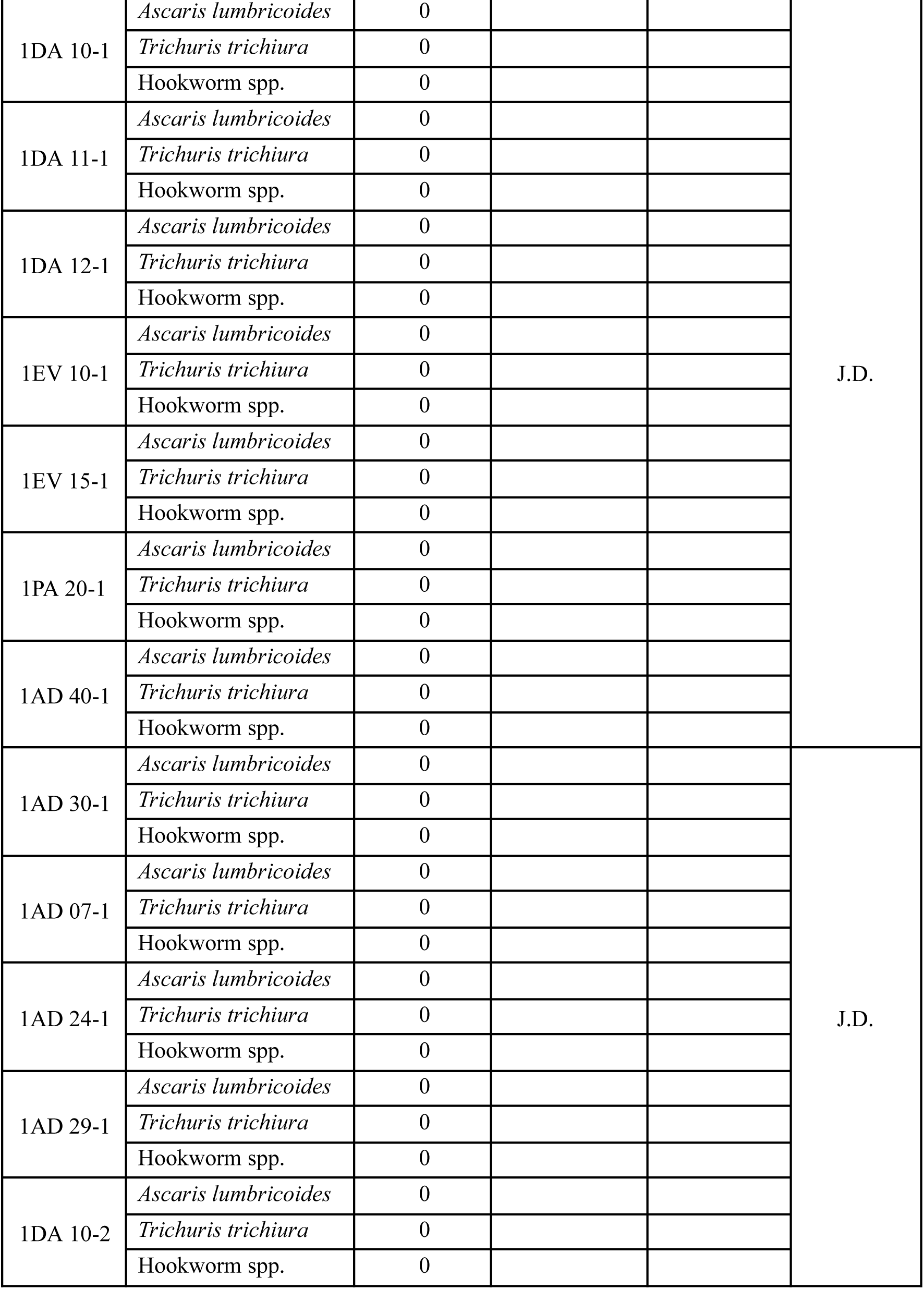
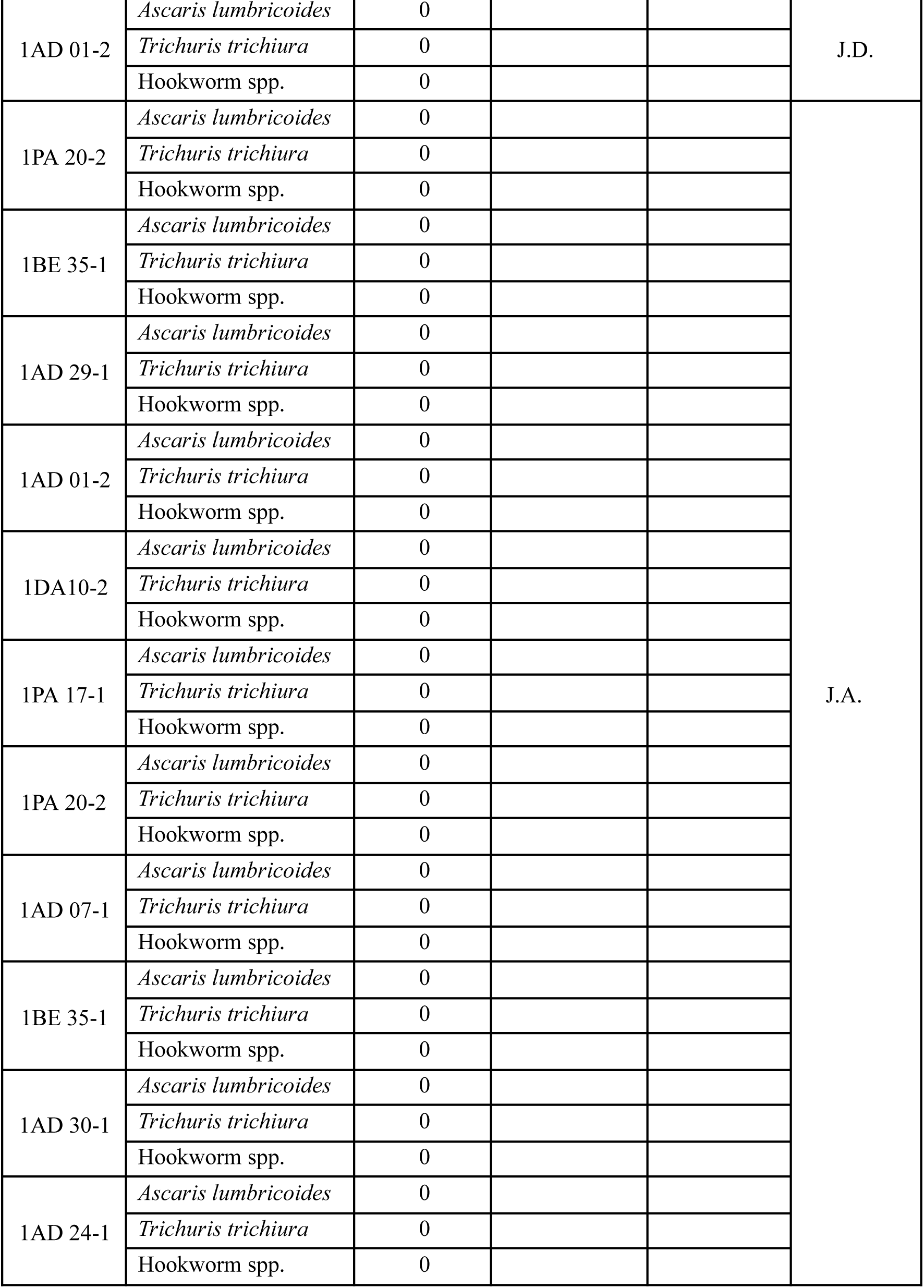
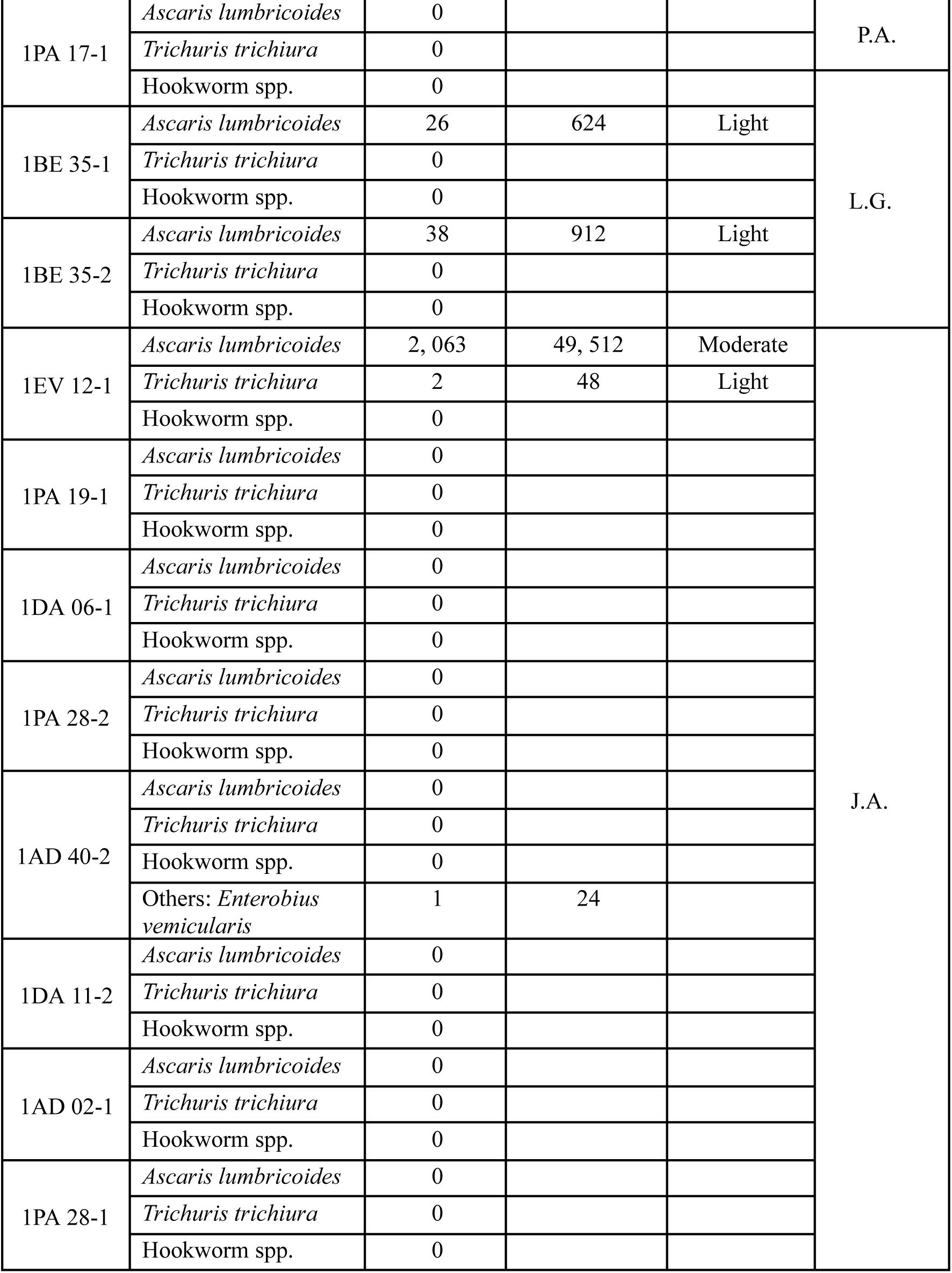
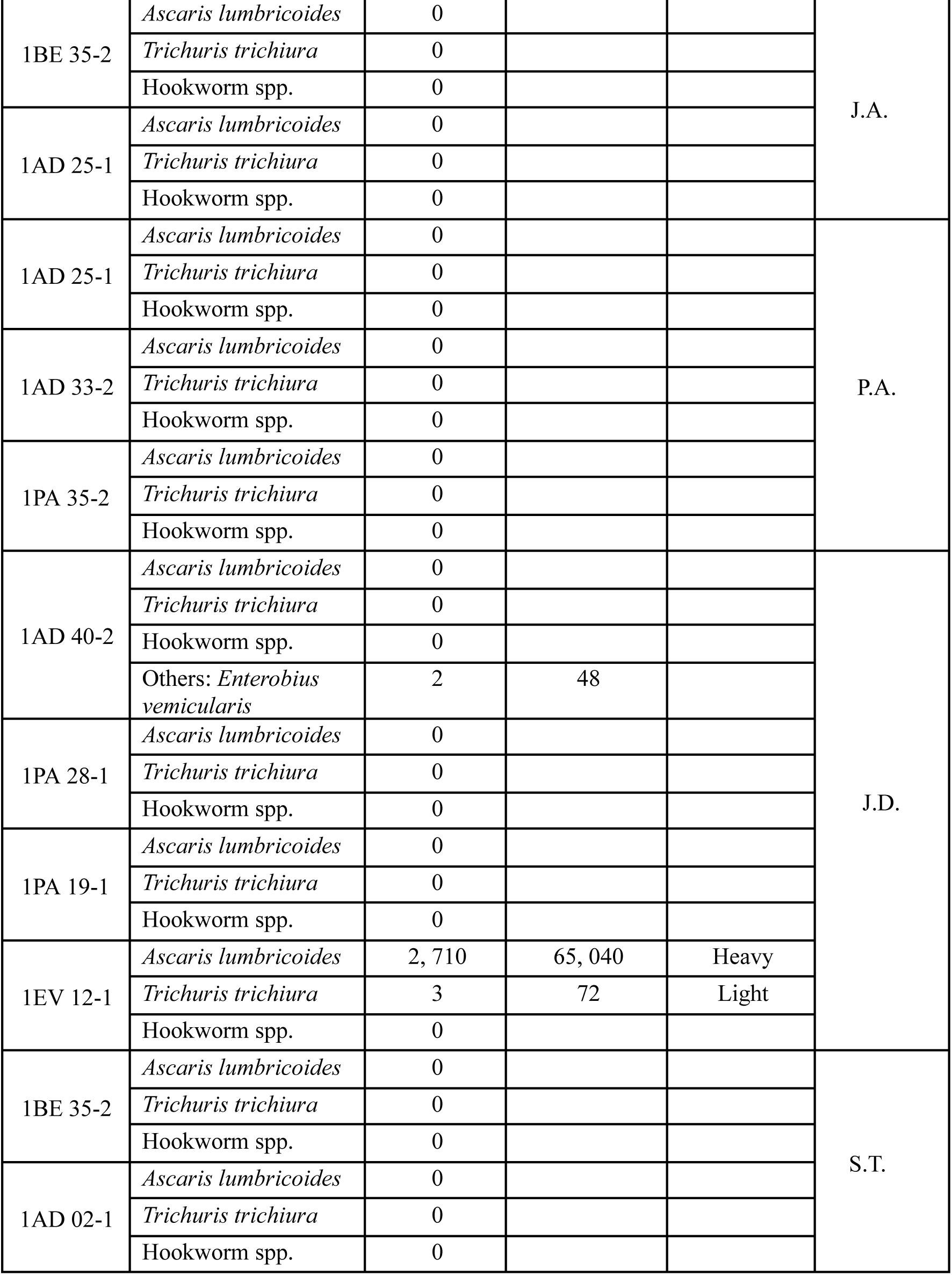
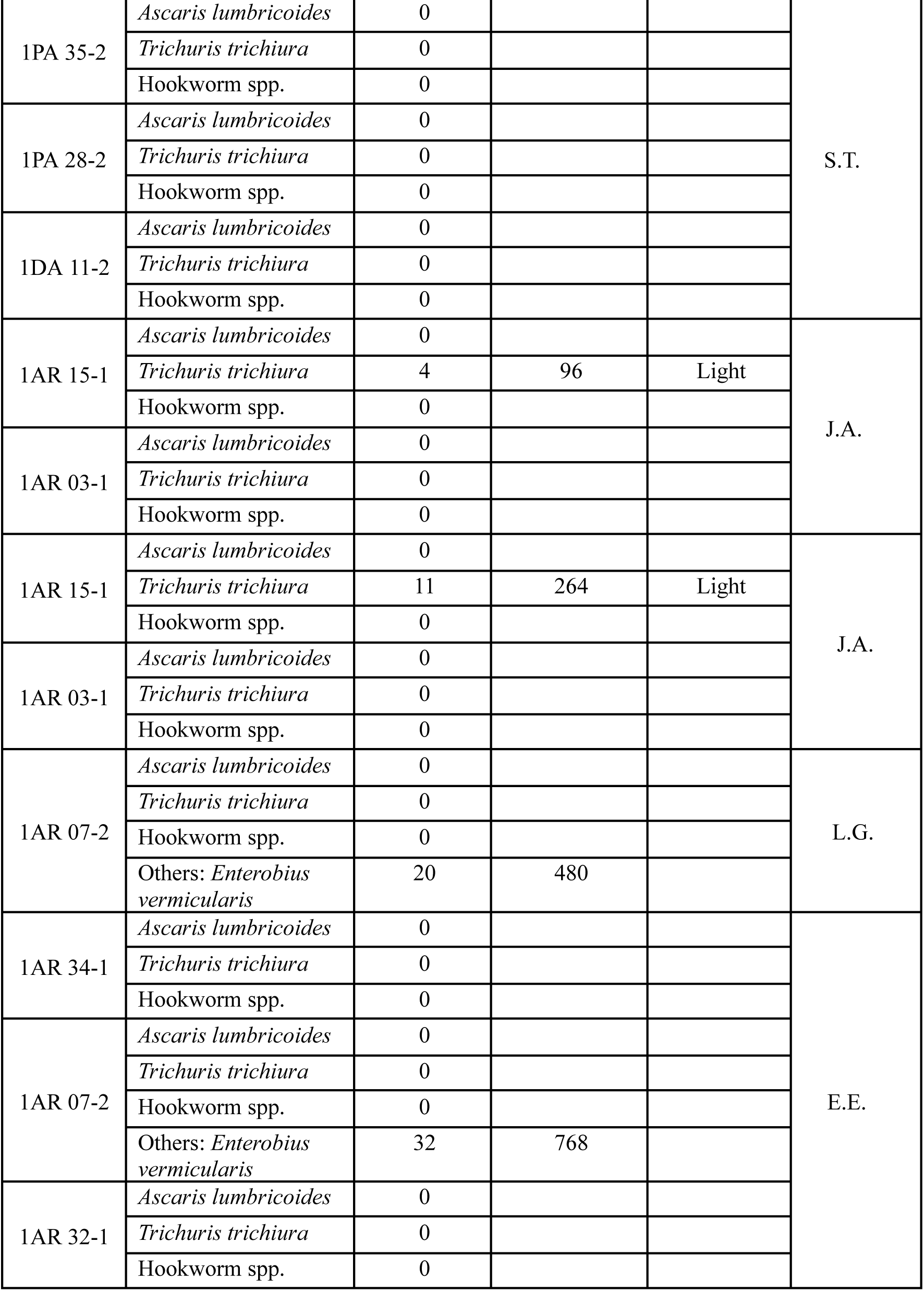
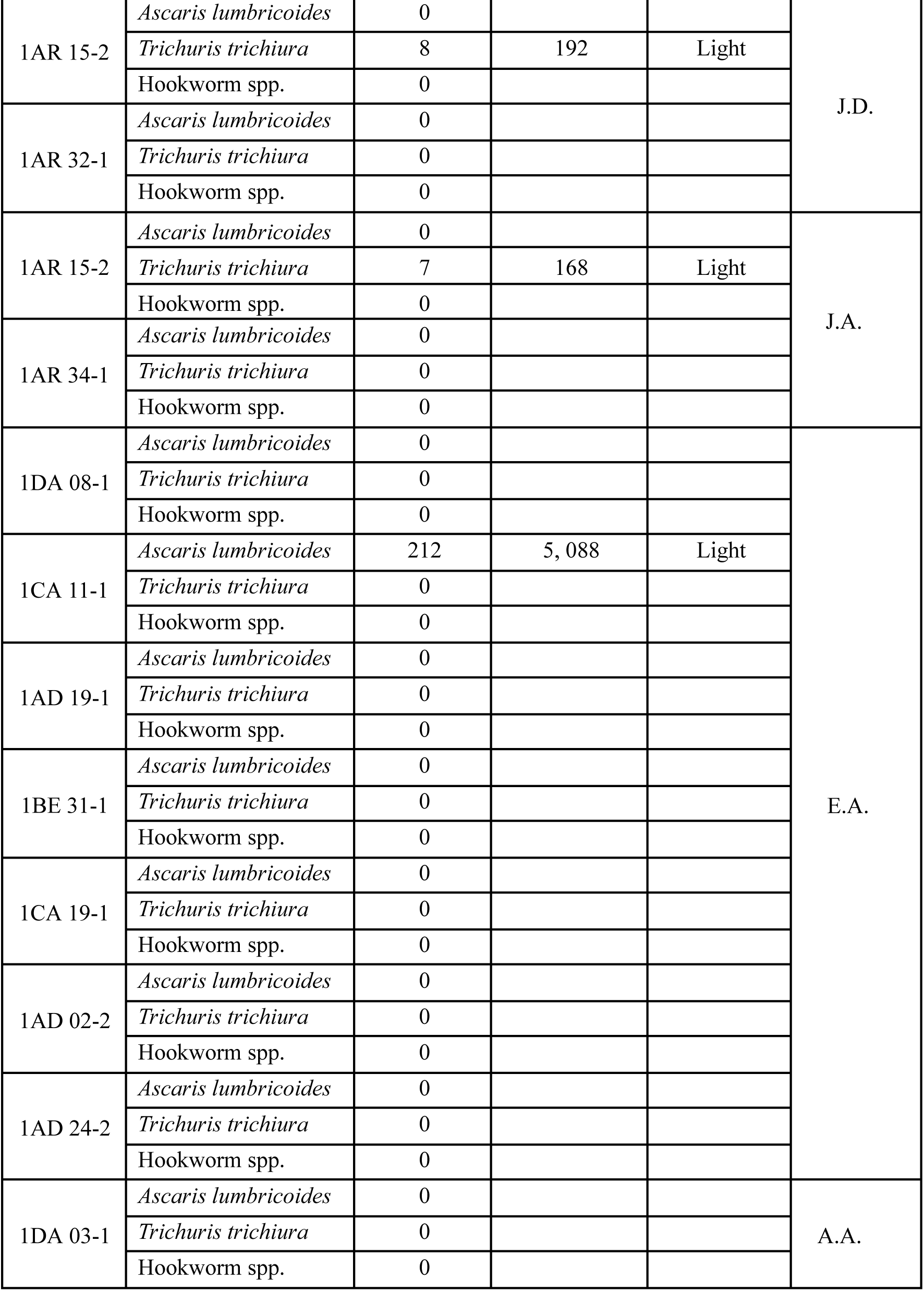
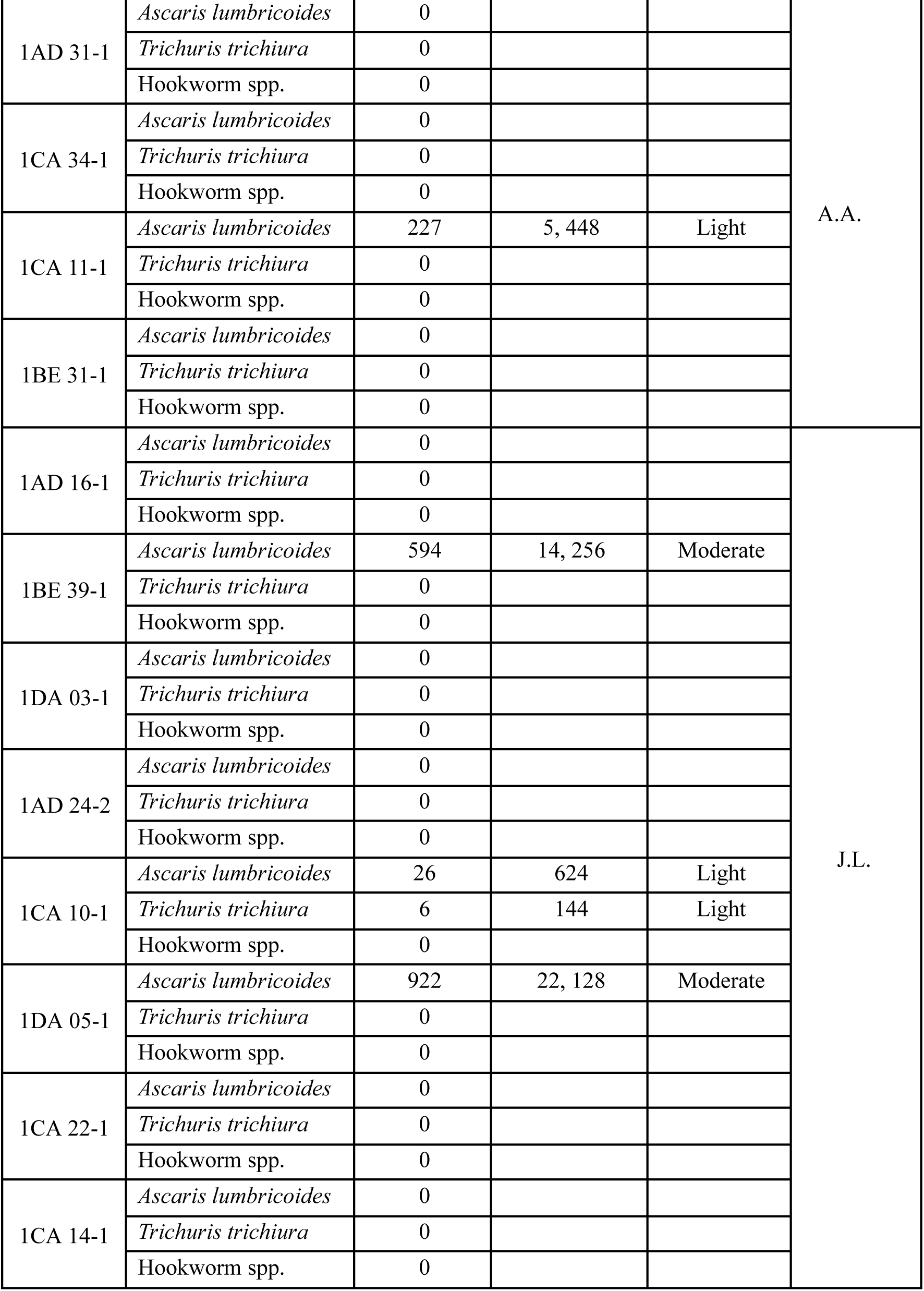
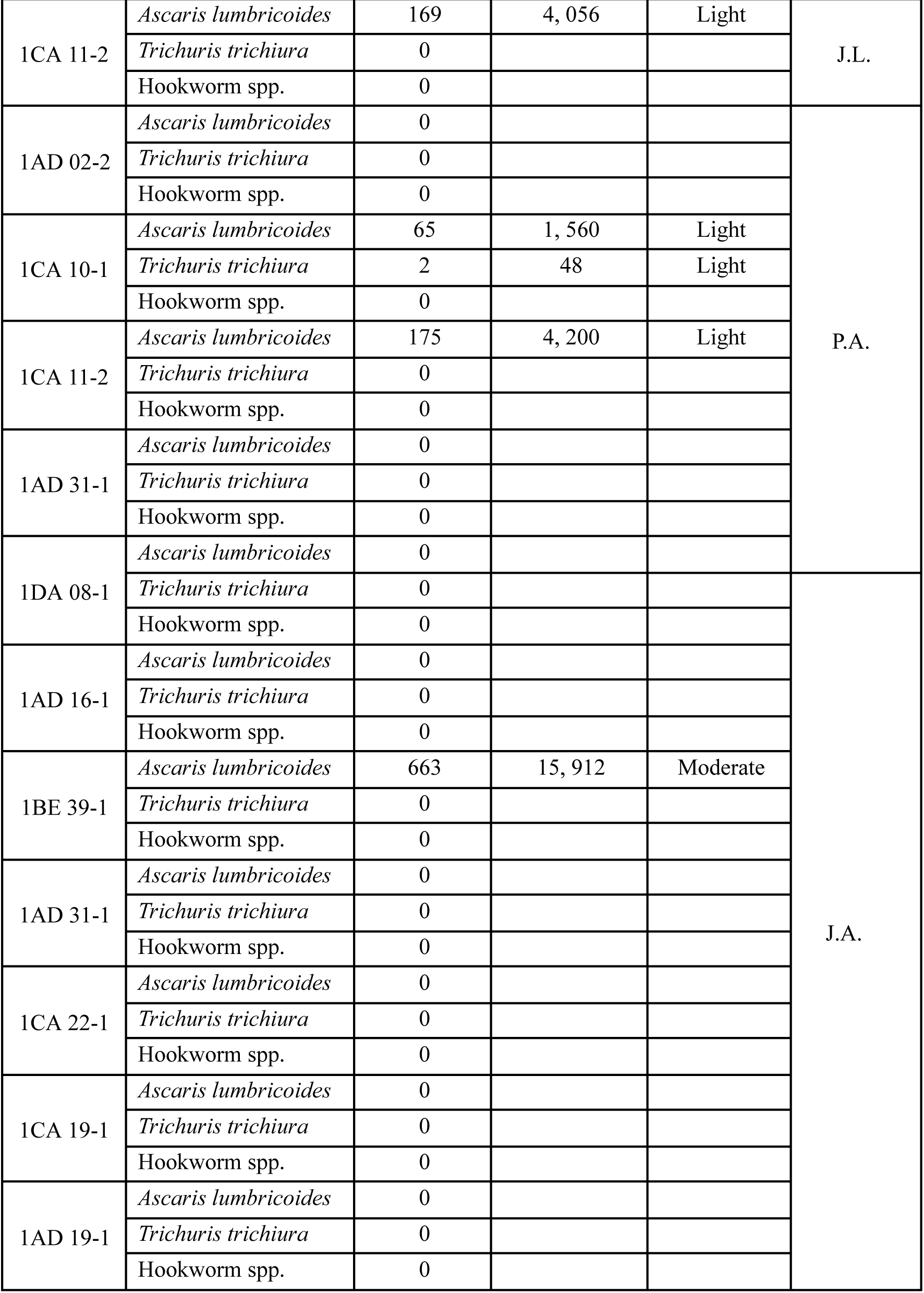
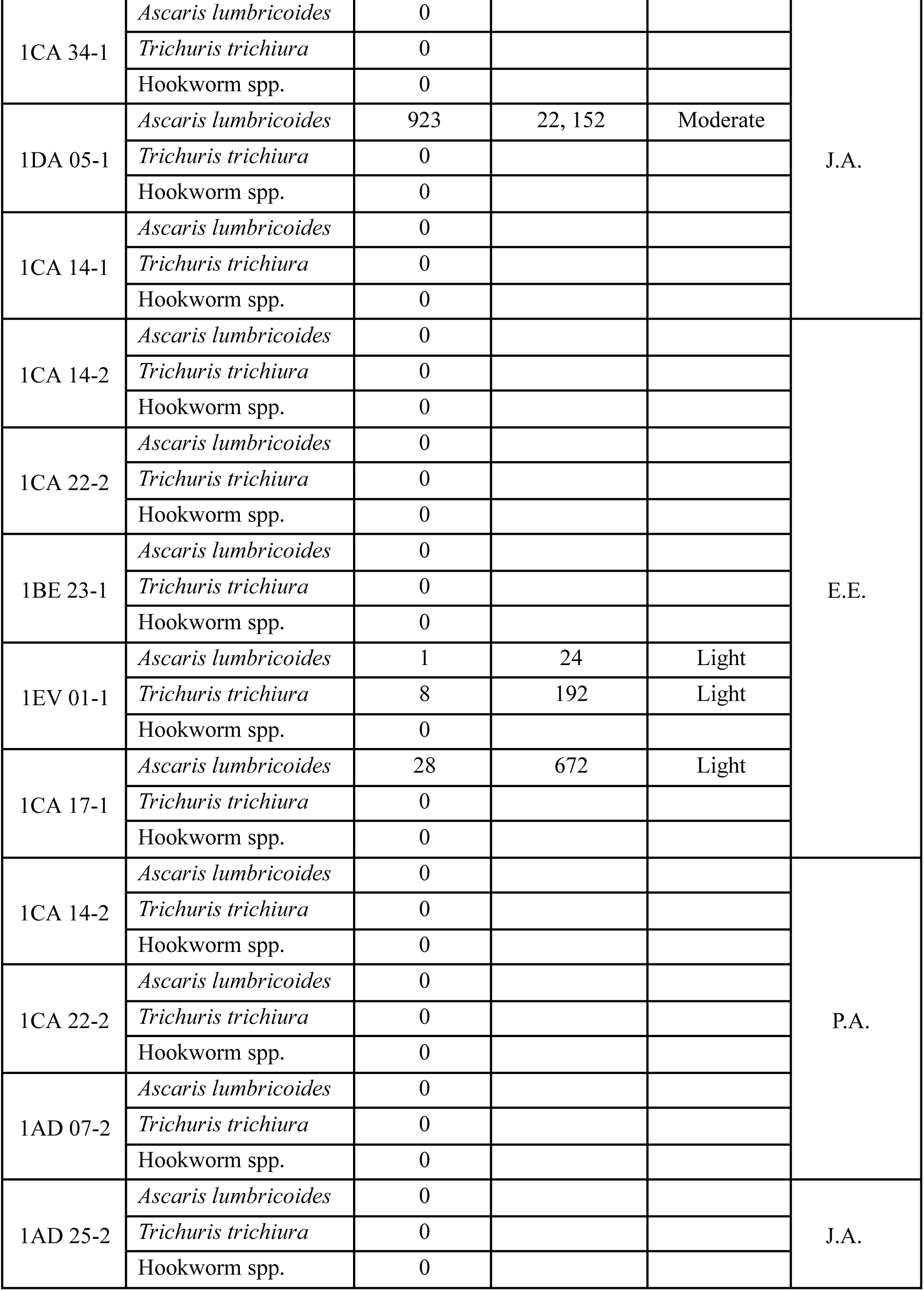
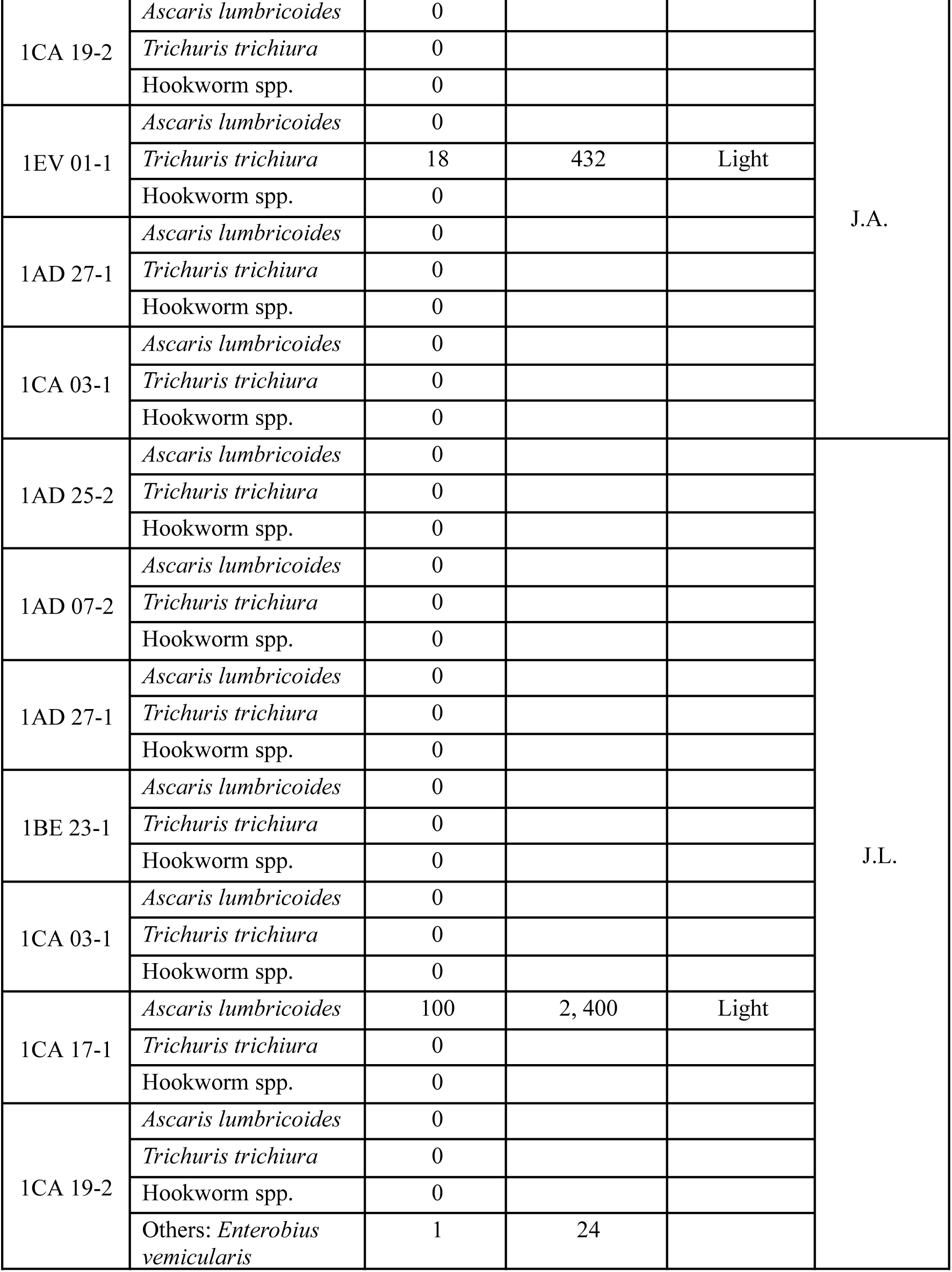
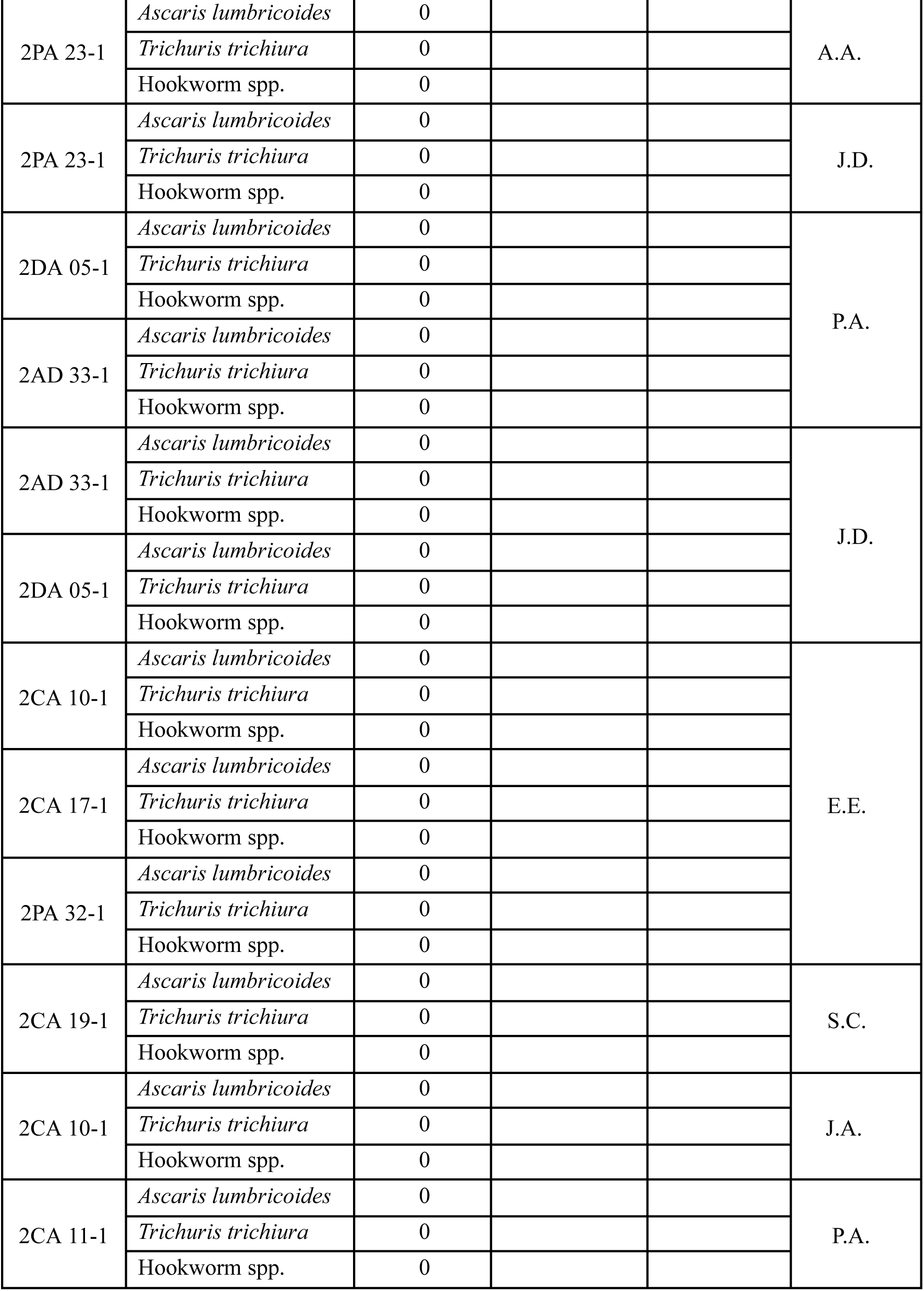
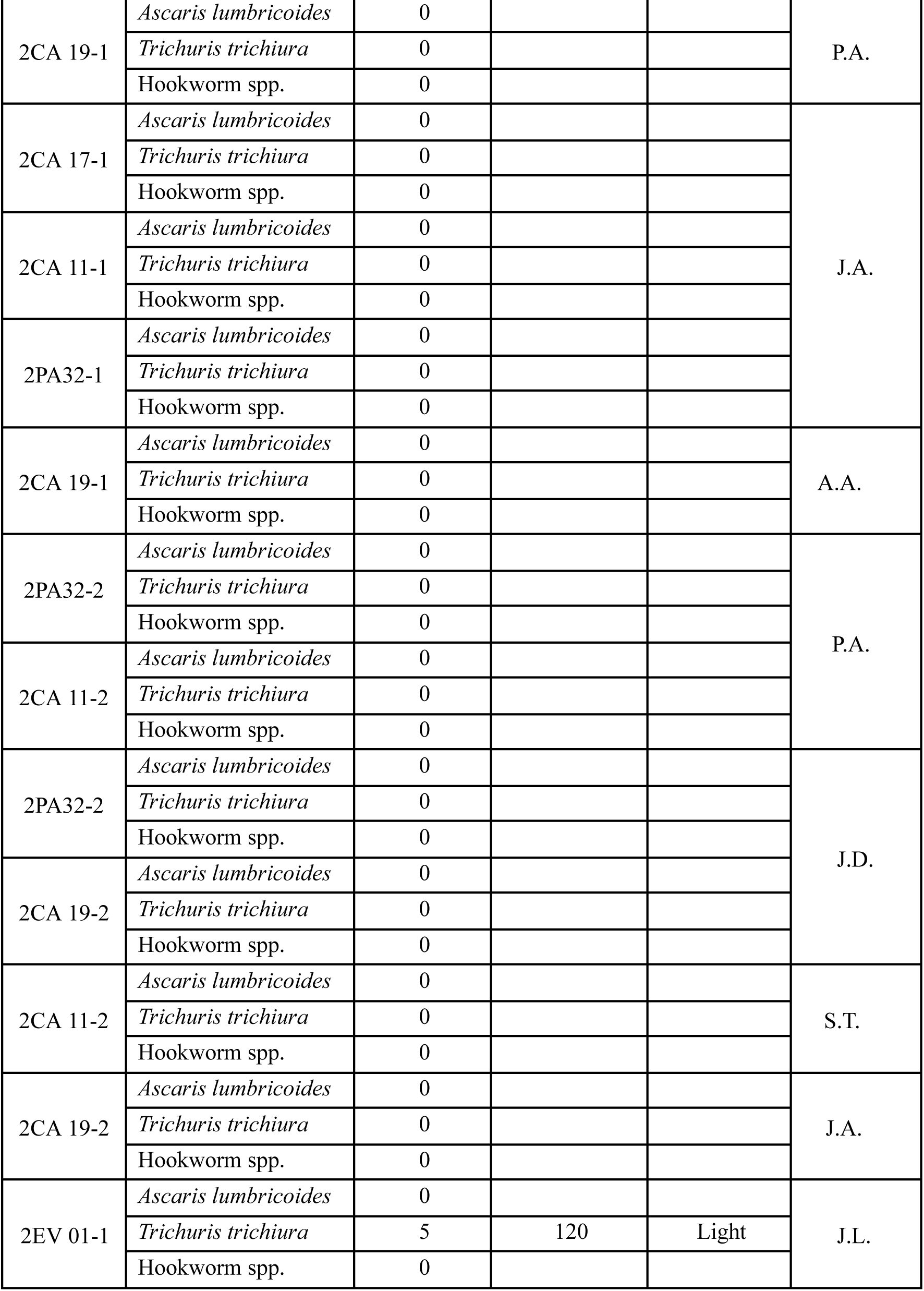
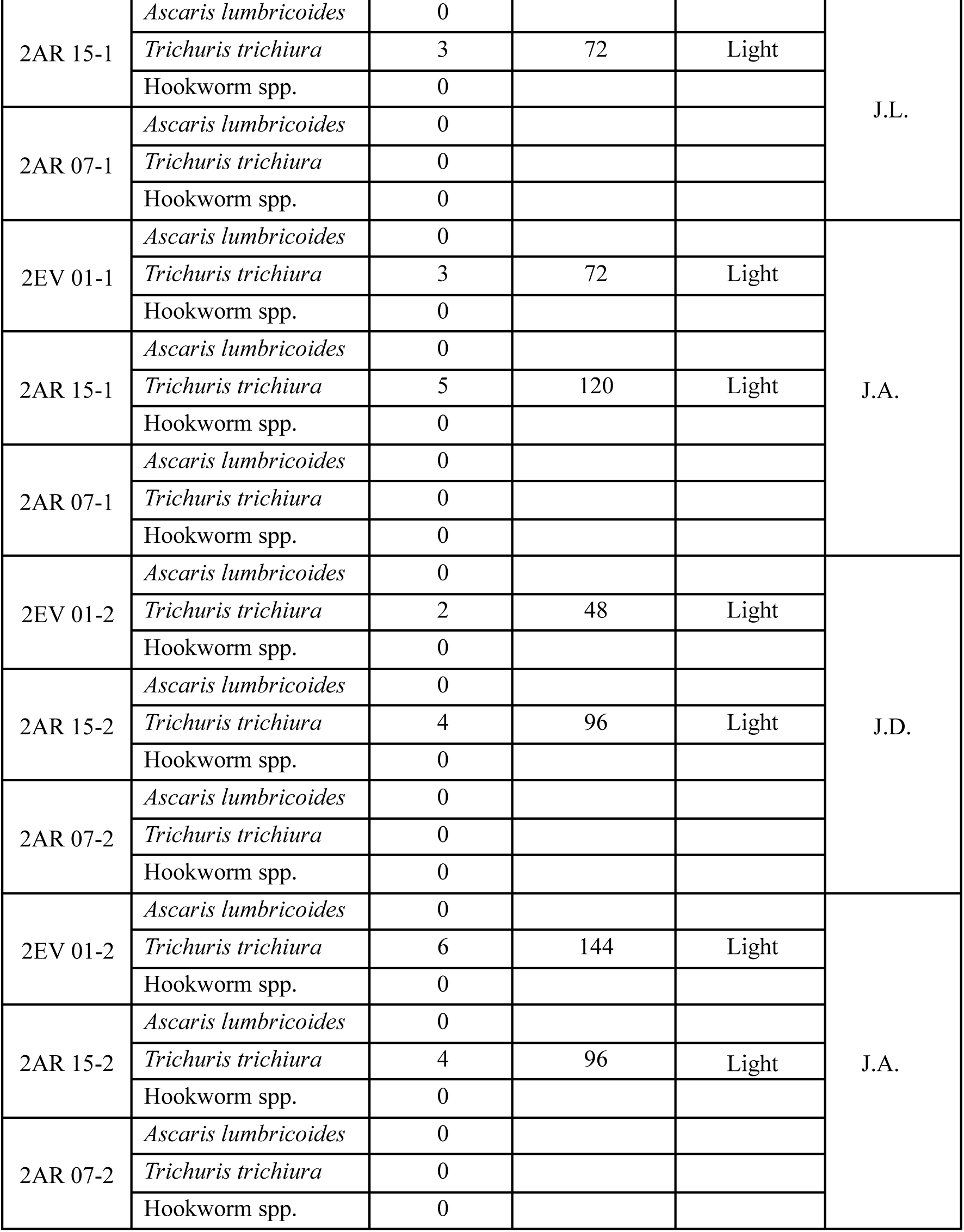

## Appendix C INFORMED CONSENT AND ASSENT FORM FOR GRADE FIVE STUDENTS

This form is for the parent/guardian and students of grade five, male and female, who are currently enrolled in Elementary School and who we are inviting to participate in the research study entitled: “The Effectiveness of Albendazole in Reducing the Prevalence and Intensity of Soil-Transmitted Helminth Infection among 9 to 12 year-old students from 5^th^ Grade of a Public Elementary School in the Philippines”

### PART I: INFORMATION SHEET

#### INTRODUCTION

We are second year students of Cebu Institute of Medicine who are conducting a research study regarding the effectiveness of the drug Albendazole on the following soil transmitted helminths: a) Ascaris lumbricoides, b) Trichuris trichiura, and c) Hookworm spp.

#### PURPOSE

We want to know the effectiveness of the drug, Albendazole, on the prevalence of soil-transmitted helminths among the students of this Elementary School because we want to provide the community with information that might help improve its promotion of health

#### PROCEDURES

If you choose to participate in this study, these following events will happen:

- You will be asked to provide two fecal specimens in clean specimen containers twice:

1. Before the deworming in July 2019 and
2. 14 – 21 days after taking the deworming medication (for those who will be positive for soil transmitted helminths)
- You will be given an orientation about the proper way of collecting the stool specimen.
- There will be no harm done to you. The collection of specimen will be done at your respective houses to ensure safety.
- Refreshments will be given to you to thank you for your participation in the study.
- The stool specimens will be collected by members of the research team and brought to and examined in the Microbiology laboratory of Cebu Institute of Medicine.

I have checked with the child and he/she understands the procedures.

*_____________________________________*

(Signature of parent/guardian)

#### VOLUNTARY PARTICIPATION

We are inviting you to be a part of this research study, but it is up to you to choose whether or not to participate. This information sheet will explain to your parent/guardian that we are asking for your agreement. If you are willing to join, your parent/guardian also have to agree. If you do not want to take part in this research, you do not have to, even if your parent/ guardian has/have agreed. I have checked with the child and he/she understands that participation is voluntary. I have checked with the child and he/she understands the procedures.

*_____________________________________*

(Signature of parent/guardian)

#### CONFIDENTIALITY

Instead of your name, a control number will be assigned to any information about you to ensure your privacy. Only the researchers will know your assigned control number.

#### SHARING OF FINDINGS

If your stool specimen comes out positive for any of the soil transmitted helminths, it will be shared with you. If you have any concerns about any part of the study or your results, please don’t hesitate to contact Joan Dihiansan (Research leader) at 09228202078.

### PART II: CERTIFICATE OF CONSENT

I have read this information (or had the information read to me). I have had my questions answered and know that I can ask questions later if I have them.

**For the Parent / Guardian:**

▭ I will allow my child, to take part in the research study.
▭ I will not allow my child to take part in the research study.

## Appendix D

Regional Director

DOH – CHD7

Osmeña Blvd., Cebu City

We are first year students from the Cebu Institute of Medicine. In partial fulfillment of the requirements for first year in Medicine, we are conducting a study entitled *The Effectiveness of Albendazole in Reducing the Prevalence and Intensity of Soil-Transmitted Helminth Infection among 9 to 12 year-old students from 5^th^ Grade of a Public Elementary School in the Philippines*. This study seeks to address the uprising cases of soil helminthiasis by comparing the baseline and follow-up prevalence and intensity of STH infections among the respondents, and determining the presence of a correlation between certain associated risk factors with the incidence of the infection. We believe that this study will be of great use not just for the well-being of the community, but also for the Health Sector and future researchers so that a clearer understanding on the nature of the disease can be established.

Though our knowledge on the topic at hand is taken from literature and text, it still is not suffice to answer all our queries and concerns. We believe that years of experience in the field is unparalleled, which is why we would like to ask your humble office for an appointment with whoever may able to share their knowledge and expertise regarding the Deworming Program and its many aspects – the processes done, the drugs given and the many technicalities to it. Aside from that we would like to ask permission to gain access to the list of schools participating in the Deworming Program, if any. It would be beneficial for our study if we can gain access to these data only while upholding the proper ethical considerations and decorum.

Please do consider our fervent request. Truly it would be a great honor to be able to partner with the Department of Health on this study. A response by April 17, 2019 would greatly be appreciated. Would you have any queries or concerns regarding the study, you may contact the cellphone number written below.

Respectfully yours,

Joan Dihiansan

*Principal Investigator*

*0922 820 2078*

Schools Division Superintendent

Dep-Ed Region 7

Imus Avenue, Cebu City

Greetings of peace and goodwill!

We are a group of first year students from the Cebu Institute of Medicine. We are conducting a research project entitled *The Effectiveness of Albendazole in Reducing the Prevalence and Intensity of Soil-Transmitted Helminth Infection among 9 to 12 year-old students from 5^th^ Grade of a Public Elementary School in the Philippines.* This study aims to compare the baseline and follow-up prevalence and intensity of STH infections among the respondents, and determine the presence of a correlation between certain associated risk factors with the incidence of the infection. We believe that this study will be of great use not just for the well-being of the community, but also for the Health Sector and future researchers so that a clearer understanding on the nature of the disease can be established.

Vital to our study is the participation of students in one school and in this regards we are inviting you to participate in this project. This is why we would like to ask for your permission to gain access and, thus, conduct our study in this Elementary School. The said pupils that shall take part in our study will undergo Fecal Examination using the Kato-Katz Technique before and after they undergo the Deworming process. As researchers, we are aware of the requisites this study will entail especially on the students. Their safety and well-being will be kept a top priority for the entire duration of this study. An Informed Consent will be distributed to each participating student’s guardian to ensure transparency. We are yet on the pre-experimentation phase and are open to discuss with your office any matter for clarification or concerns.

Please do consider our fervent request. Truly it would be a great honor to be able to partner with the Department of Education and the Department of Health on this study. We appreciate if your office can attend to this as soon as possible. Would you have any queries or concerns regarding the study, you may contact the cellphone number written below.

Respectfully yours,

Joan Dihiansan

*Principal Investigator*

*0922 820 2078/0917 126 0979*

Dr.

Year II Coordinator

Cebu Institute of Medicine

Good day!

We are a group of first year students from the Cebu Institute of Medicine. We are conducting a research project entitled *The Effectiveness of Albendazole in Reducing the Prevalence and Intensity of Soil-Transmitted Helminth Infection among 9 to 12 year-old students from 5^th^ Grade of a Public Elementary School in the Philippines.* This study aims to compare the baseline and follow-up prevalence and intensity of STH infections among the respondents, and determine the presence of a correlation between certain associated risk factors with the incidence of the infection. We believe that this study will be of great use not just for the well-being of the community, but also for the Health Sector and future researchers so that a clearer understanding on the nature of the disease can be established.

We will be taking our stool samples from Grade 4 to 6 students of an Elementary School. The said pupils that shall take part in our study will undergo Fecal Examination using the Kato-Katz Technique before and after they undergo the Deworming process. In line with this, the group would like to ask for your permission to use the Microbiology Laboratory at the Cebu Institute of Medicine as venue for our Stool examination. The materials needed for the Kato-Katz technique will be sponsored by the Department of Health.

We hope for your kind consideration.

Respectfully yours,

Joan Dihiansan

*Principal Investigator*

*0922 820 2078/0917 126 0979*

